# Large-scale genome-wide association study of 398,238 women unveils seven novel loci associated with high-grade serous epithelial ovarian cancer risk

**DOI:** 10.1101/2024.02.29.24303243

**Authors:** Daniel R. Barnes, Jonathan P. Tyrer, Joe Dennis, Goska Leslie, Manjeet K. Bolla, Michael Lush, Amber M. Aeilts, Kristiina Aittomäki, Nadine Andrieu, Irene L. Andrulis, Hoda Anton-Culver, Adalgeir Arason, Banu K. Arun, Judith Balmaña, Elisa V. Bandera, Rosa B. Barkardottir, Lieke P.V. Berger, Amy Berrington de Gonzalez, Pascaline Berthet, Katarzyna Białkowska, Line Bjørge, Amie M. Blanco, Marinus J. Blok, Kristie A. Bobolis, Natalia V. Bogdanova, James D. Brenton, Henriett Butz, Saundra S. Buys, Maria A. Caligo, Ian Campbell, Carmen Castillo, Kathleen B.M. Claes, GEMO Study Collaborators, EMBRACE Collaborators, Sarah V. Colonna, Linda S. Cook, Mary B. Daly, Agnieszka Dansonka-Mieszkowska, Miguel de la Hoya, Anna deFazio, Allison DePersia, Yuan Chun Ding, Susan M. Domchek, Thilo Dörk, Zakaria Einbeigi, Christoph Engel, D. Gareth Evans, Lenka Foretova, Renée T. Fortner, Florentia Fostira, Maria Cristina Foti, Eitan Friedman, Megan N. Frone, Patricia A. Ganz, Aleksandra Gentry-Maharaj, Gord Glendon, Andrew K. Godwin, Anna González-Neira, Mark H. Greene, Jacek Gronwald, Aliana Guerrieri-Gonzaga, Ute Hamann, Thomas v.O. Hansen, Holly R. Harris, Jan Hauke, Florian Heitz, Frans B.L. Hogervorst, Maartje J. Hooning, John L. Hopper, Chad D Huff, David G. Huntsman, Evgeny N. Imyanitov, kConFab Investigators, Louise Izatt, Anna Jakubowska, Paul A. James, Ramunas Janavicius, Esther M. John, Siddhartha Kar, Beth Y. Karlan, Catherine J. Kennedy, Lambertus A.L.M. Kiemeney, Irene Konstantopoulou, Jolanta Kupryjanczyk, Yael Laitman, Ofer Lavie, Kate Lawrenson, Jenny Lester, Fabienne Lesueur, Carlos Lopez-Pleguezuelos, Phuong L. Mai, Siranoush Manoukian, Taymaa May, Iain A. McNeish, Usha Menon, Roger L. Milne, Francesmary Modugno, Jennifer M. Mongiovi, Marco Montagna, Kirsten B. Moysich, Susan L. Neuhausen, Finn C. Nielsen, Catherine Noguès, Edit Oláh, Olufunmilayo I. Olopade, Ana Osorio, Laura Papi, Harsh Pathak, Celeste L. Pearce, Inge S. Pedersen, Ana Peixoto, Tanja Pejovic, Pei-Chen Peng, Beth N. Peshkin, Paolo Peterlongo, C. Bethan Powell, Darya Prokofyeva, Miquel Angel Pujana, Paolo Radice, Muhammad U. Rashid, Gad Rennert, George Richenberg, Dale P. Sandler, Naoko Sasamoto, Veronica W. Setiawan, Priyanka Sharma, Weiva Sieh, Christian F. Singer, Katie Snape, Anna P. Sokolenko, Penny Soucy, Melissa C. Southey, Dominique Stoppa-Lyonnet, Rebecca Sutphen, Christian Sutter, Manuel R. Teixeira, Kathryn L. Terry, Liv Cecilie V. Thomsen, Marc Tischkowitz, Amanda E. Toland, Toon Van Gorp, Ana Vega, Digna R. Velez Edwards, Penelope M. Webb, Jeffrey N. Weitzel, Nicolas Wentzensen, Alice S. Whittemore, Stacey J. Winham, Anna H. Wu, Siddhartha Yadav, Yao Yu, Argyrios Ziogas, Andrew Berchuck, Fergus J. Couch, Ellen L. Goode, Marc T. Goodman, Alvaro N. Monteiro, Kenneth Offit, Susan J. Ramus, Harvey A. Risch, Joellen M. Schildkraut, Mads Thomassen, Jacques Simard, Douglas F. Easton, Michelle R. Jones, Georgia Chenevix-Trench, Simon A. Gayther, Antonis C. Antoniou, Paul D.P. Pharoah, the Ovarian Cancer Association Consortium and the Consortium of Investigators of Modifiers of BRCA1 and BRCA2

## Abstract

**Background:** Nineteen genomic regions have been associated with high-grade serous ovarian cancer (HGSOC). We used data from the Ovarian Cancer Association Consortium (OCAC), Consortium of Investigators of Modifiers of *BRCA1*/*BRCA2* (CIMBA), UK Biobank (UKBB), and FinnGen to identify novel HGSOC susceptibility loci and develop polygenic scores (PGS).

**Methods:** We analyzed >22 million variants for 398,238 women. Associations were assessed separately by consortium and meta-analysed. OCAC and CIMBA data were used to develop PGS which were trained on FinnGen data and validated in UKBB and BioBank Japan

**Results:** Eight novel variants were associated with HGSOC risk. An interesting discovery biologically was finding that *TP53* 3’-UTR SNP rs78378222 was associated with HGSOC (per T allele relative risk (RR)=1.44, 95%CI:1.28-1.62, P=1.76×10^-9^). The optimal PGS included 64,518 variants and was associated with an odds ratio of 1.46 (95%CI:1.37-1.54) per standard deviation in the UKBB validation (AUROC curve=0.61, 95%CI:0.59-0.62).

**Conclusions:** This study represents the largest GWAS for HGSOC to date. The results highlight that improvements in imputation reference panels and increased sample sizes can identify HGSOC associated variants that previously went undetected, resulting in improved PGS. The use of updated PGS in cancer risk prediction algorithms will then improve personalized risk prediction for HGSOC.

## INTRODUCTION

Globally, epithelial ovarian cancer (EOC) is the seventh most common cancer diagnosed in women, with approximately 314,000 new cases diagnosed each year^1^. It is the most lethal gynecological cancer, responsible for approximately 207,000 deaths annually^1^. EOC is a collection of five major histotypes, namely high-grade serous (HGSOC), endometrioid, clear cell, low-grade serous (LGS) and mucinous, which are thought to have distinct etiology^2^. HGSOC is the most prevalent accounting for 60-70% of EOC diagnoses^2,3^, and accounting for most EOCs diagnosed in *BRCA1* and *BRCA2* pathogenic variant (PV) carriers^4–11^. Furthermore, HGSOC accounts for the majority of EOC mortality^12,13^.

To date, 40 genomic regions associated with EOC have been identified through genome-wide association studies (GWAS)^14–27^[OCAC fine-mapping article, accepted AJHG]. For 19 of these regions, HGSOC is the most strongly associated histotype^14–22,26,27^[OCAC fine-mapping article, accepted AJHG]. These studies have relied on imputation efforts that used the 1000 Genomes Project^28^ and Haplotype Reference Consortium^29^ reference panels, yielding up to approximately 11 million well-imputed genetic variants. The Trans-Omics for Precision Medicine (TOPMed) reference panel^30^ and imputation server^31^ have recently become publicly available. The TOPMed panel consists of approximately 308 million variants, yielding greater genomic coverage than previously available reference panels, with the added benefit of containing many more low-frequency and rare variants. This prompted us to re-impute genetic variant data from the population-based Ovarian Cancer Association Consortium (OCAC)^32^, and *BRCA1*/*2* carriers from the Consortium of Investigators of Modifiers of *BRCA1* and *BRCA2* (CIMBA)^33,34^ to assess whether the larger coverage of the genome from the TOPMed reference panel leads to detection of novel loci associated with HGSOC risk. We additionally made use of the UK Biobank (UKBB)^35,36^ to boost the sample size and power to detect associations. We combined these data with summary statistics from FinnGen^37,38^ and BioBank Japan^39,40^ to develop and validate polygenic models (PGM) and scores (PGS) for non-mucinous OC.

## METHODS

### Study samples

OCAC participants were enrolled in 65 studies from 16 countries and a large European multinational nested case-control study (Supplementary Table 1). OCAC individual participant data were used for GWAS discovery analyses and developing polygenic models (PGMs).

CIMBA study participants were enrolled in 64 studies from 28 countries (Supplementary Table 2). Eligibility was restricted to women aged at least 18 years at the time of recruitment who carried a PV in either *BRCA1* or *BRCA2*. Data collected included year of birth, PV description, age at recruitment, age at last follow-up, and age at breast and ovarian cancer (invasive, fallopian tube and peritoneal) diagnoses, and age or date of prophylactic surgeries (bilateral mastectomy and bilateral oophorectomy). Most participants were recruited through cancer genetics clinics and enrolled in regional/national research studies. CIMBA individual participant data were used in the GWAS discovery and in PGM development.

The UK Biobank (UKBB) is a large-scale biomedical research resource, with detailed genetic and health data on half a million UK participants^35,36^. For the purposes of these analyses, data from 245,377 female participants of European ancestry were used. UKBB individual participant data were used in the GWAS discovery analyses and to independently validate PGS.

FinnGen is a large collection of newly recruited and legacy samples from Finnish biobanks, research institutes, universities, university hospitals, international pharmaceutical partners, the Finnish Blood Service, the Finnish Biobank Cooperative, and the Finnish Institute for Health and Welfare, utilizing Finnish longitudinal health register data collected on every resident of Finland since 1969^37,38^. FinnGen summary statistics based on 150,658 women (149,394 controls, 1,264 EOC cases of any histotype) were used to train PGM hyperparameters.

BioBank Japan (BBJ) is a large biobank resource containing clinical and genetic data on over 300,000 participants^39,40^. BBJ summary statistics based on 61,457 women (60,614 controls, 843 EOC cases) were used for assessing PGS associations for women of East Asian ancestry.

### Genotyping and re-imputation using the TOPMed reference panel

Genotyping for OCAC and CIMBA was performed on one of two custom single nucleotide polymorphism (SNP) arrays (iCOGS and OncoArray), described in the Supplementary Methods. OCAC had additional samples genotyped on GWAS arrays^14–16^. These data were imputed to the TOPMed reference panel^30^ after standard quality control (Supplementary Methods). Details of the UKBB genotyping and imputation to a combined UK10K^41,42^ and HRC^29^ reference panel have been described elsewhere^35,36^. The OCAC, CIMBA and UKBB analyses were based on 142 million, 104 million and 60 million well-imputed (imputation *r*^2^>0.30) variants, respectively (Table 1). Downstream meta-analyses were restricted to variants that had minor allele counts (MACs) of MAC>5 and did not have heterogeneous effects (P_het_>1×10^-8^) in the meta-analysis of OCAC studies. Analyses including FinnGen and BBJ data made use of summary statistic data only. Details of FinnGen and BBJ genotyping and imputation have been described elsewhere^37–40^.

### Statistical analyses of OCAC and UKBB data

We examined the associations between genotypes and HGSOC risk in the OCAC data using logistic regression. Analyses were conducted separately for OncoArray, iCOGS, and five GWAS datasets and were combined by fixed-effects inverse-variance weighted meta-analysis (Figure 1). We included project-specific principal components (PCs) as covariates in the model with the number of PCs based on the inflection point observed in a scree plot. All women were of European ancestry, determined using genetic data (Supplementary Methods)^22,43^.

**Figure 1:**
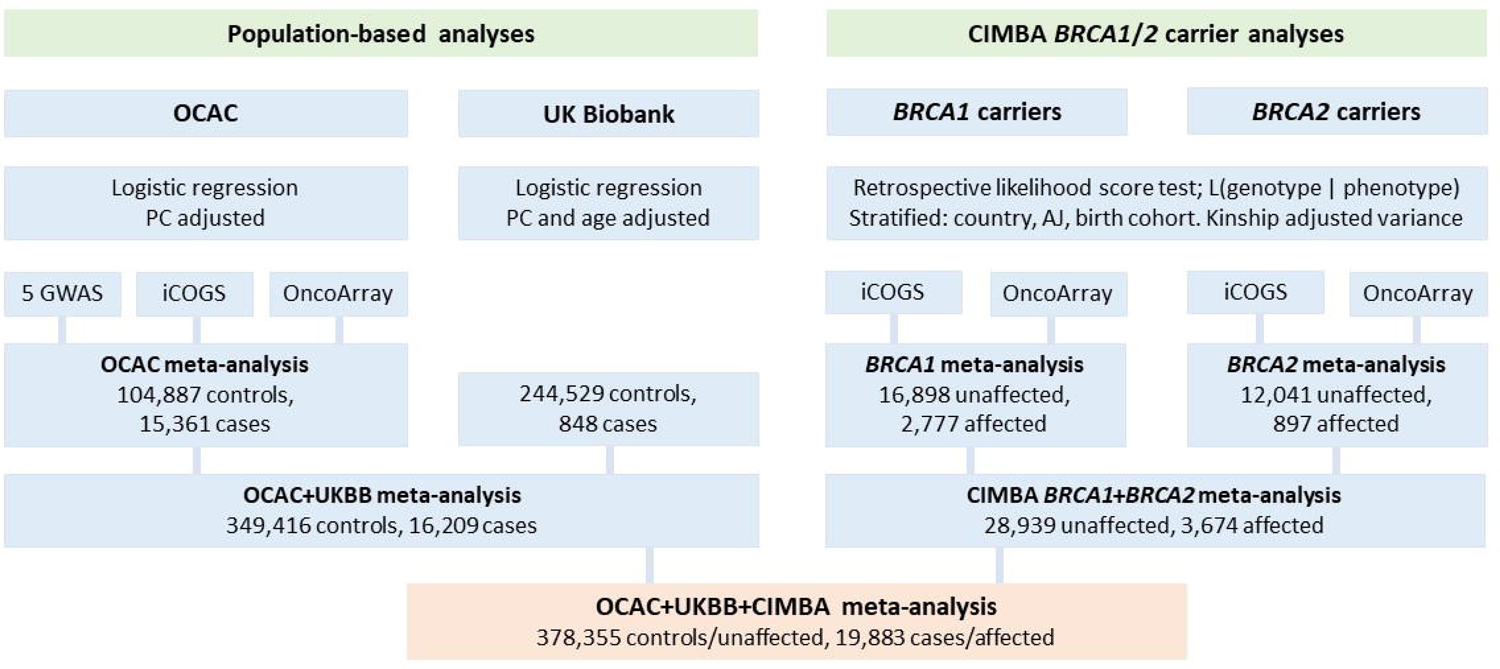
Schema describing the analytical process for the GWAS analyses from OCAC, UKBB and CIMBA, and subsequent meta-analyses.

The UKBB analysis was restricted to women with European ancestry, selected based on their PCs. EOC histotypes were classified using diagnostic codes provided by UKBB, with “serous”, “undifferentiated”, or “other” classified as HGSOC, a methodology similar to that used in OCAC^22^. Association analyses between genotypes and HGSOC risk were assessed by logistic regression (Figure 1). We adjusted for the top four PCs and age at recruitment.

### Statistical analyses of BRCA1 and BRCA2 pathogenic variant carriers

Analyses of CIMBA data were limited to carriers of European ancestry, determined by genetic data and multidimensional scaling (Supplementary Methods)^22,43^. Association analyses were performed separately by genotyping array (iCOGS or OncoArray), and separately for *BRCA1* and *BRCA2* PV carriers (Figure 1). iCOGS and OncoArray associations were combined by fixed-effects inverse-variance weighted meta-analysis to estimate *BRCA1* and *BRCA2* PV carrier specific associations. The association analysis was carried out within a survival analysis framework, by modelling the retrospective likelihood of observing the genotypes conditional on the disease phenotypes to adjust for the non-random ascertainment with respect to disease phenotypes^44,45^. The censoring process followed carriers from birth until the first occurrence of: EOC (including fallopian tube and peritoneal cancers) diagnosis, risk-reducing salpingo-oophorectomy, or study entry. Breast cancer diagnoses were not considered to be a censoring event and EOC was the endpoint of interest. Associations were then assessed using the score test statistic based on the retrospective likelihood^44,45^ assuming *BRCA1* and *BRCA2* PV carrier specific and age-cohort specific EOC incidences^46^. Analyses were stratified by country and Ashkenazi Jewish ancestry, and to account for relatedness between individuals we calculated kinship adjusted variances^47^. Only variants that were available through OncoArray genotyping and imputation were considered, as the majority of samples were available from this genotyping platform (Table 1). As HGSOC is the predominant histotype for both *BRCA1* and *BRCA2* PV carriers, the associations were combined by fixed-effects and inverse variance weighted meta-analysis using the METAL software^48^.

### Meta-analyses

We pooled the combined OCAC and UKBB summary association data (per-allele odds ratios, ORs) with the combined *BRCA1* and *BRCA2* PV carrier summary association data (per-allele hazard ratios, HRs) by fixed-effects inverse-variance weighted meta-analysis using METAL^48^ to give per-allele relative risks (RRs, a combination of population-based ORs and *BRCA1*/*2* carrier HRs) (Figure 1).

### Eliminating likely statistical artifacts

The associations of all variants with genome-wide significant associations and falling outside known regions were re-evaluated to eliminate likely spurious associations potentially due to unstable effect estimates from strata with small numbers. For this purpose, the associations were re-analysed, pooling individual level data from OCAC and UKBB (Supplementary Methods), whilst the associations for *BRCA1*/*2* PV carriers were reassessed assuming all study participants came from a single stratum.

We also re-evaluated potentially novel associations with variants in regions proximal to known regions by performing approximate conditional analysis^49^. This approach utilized summary statistics from the combined OCAC, UKBB and *BRCA1/2* PV carrier meta-analysis and the linkage disequilibrium (LD) structure from 111,304 women genotyped on the OncoArray from OCAC and CIMBA.

For variants passing these checks, we calculated Bayes false discovery probabilities (BFDPs), assuming prior probabilities of 1:1,000 and 1:10,000 variants being truly associated^50^.

### Defining credible causal variants

The sentinel variant (variant with the smallest P-value) at each novel region may not be causal. Therefore, we identified lists of credible causal variants (CCVs) that are likely to contain the genetic variant responsible for altering HGSOC risk defined as the set of variants within ±500kb of the sentinel variant whose likelihood was within a 100-fold range of the sentinel variant’s likelihood^51^.

### Development of polygenic risk models

We developed several PGMs (sets of variants and their weights) for non-mucinous OC using the Select and Shrink with Summary Statistics (S4) method^52,53^, using P-value to LD *r*^2^ ratios 0.02 and 0.15 to select variants, and each model taking different combinations of model hyperparameters (Figure 2, Supplementary Methods).

**Figure 2:**
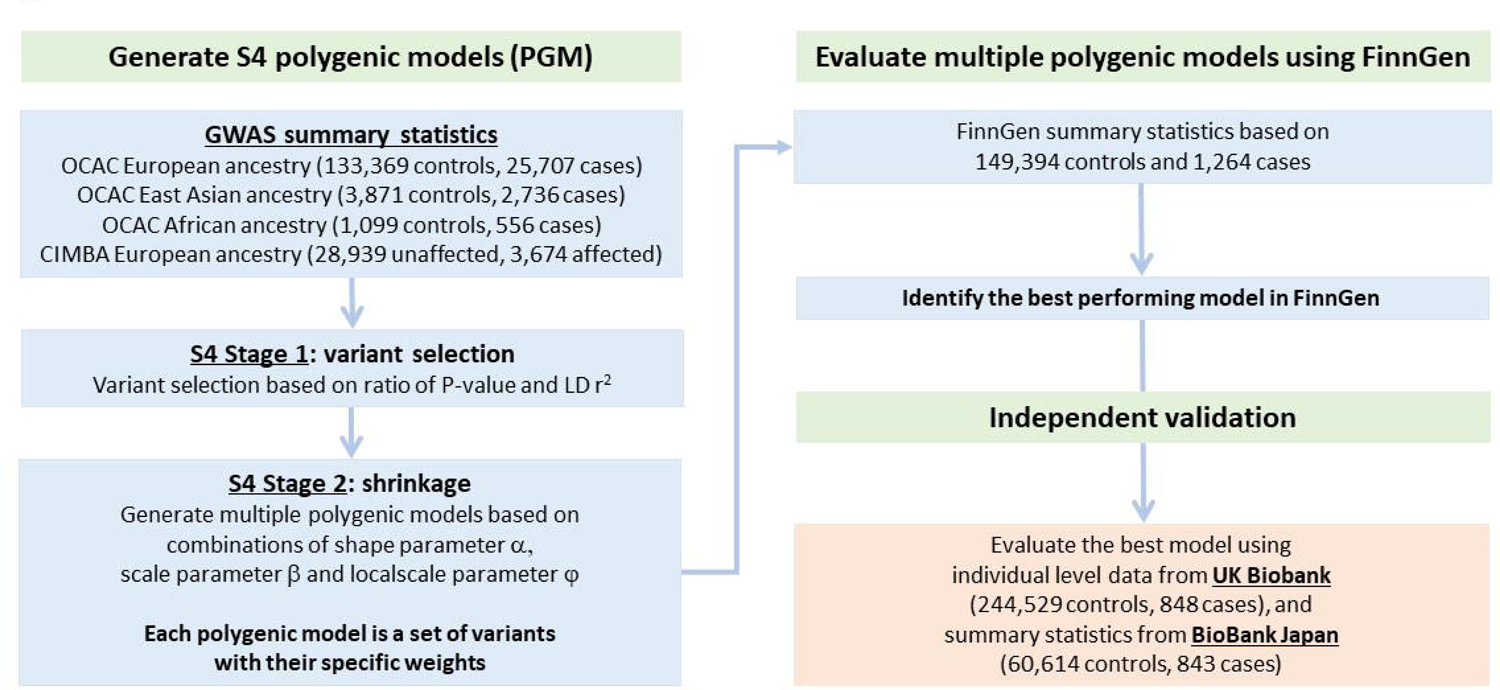
Schema describing the development of polygenic models, determining the optimal model, and validating the resultant polygenic score in European ancestry women from UKBB and East Asian ancestry women from BBJ.

Polygenic scores (PGS) are PGMs applied to observed or imputed genotypes. Previous analyses of PGMs specific to HGS, LGS, and mucinous histotypes showed that all histotypes, except mucinous, were associated with either the HGSOC PGS or LGSOC PGS. Thus, the non-mucinous histotypes were combined here^53^. The PGM was developed on summary statistics using associations obtained from a meta-analysis of the OCAC and CIMBA data. For this purpose, to maximize sample size and genetic diversity, the OCAC summary statistics were based on a meta-analysis of associations from East Asian (3,871 controls, 2,736 cases), African (1,099 controls, 556 cases), and European (133,369 controls, 25,707 cases) populations.

The LD structure was weighted to reflect the average effects from each ancestry. Model hyperparameters (shrinkage parameters controlling small and large variant effect sizes, and an overall shrinkage parameter) were trained using summary association statistics based on 150,658 women (149,394 controls, 1,264 EOC cases) from FinnGen (release 8)^37,38^. EOC histotype data was not available for FinnGen, hence we used the associations with overall EOC for the PGM training. Finally, we used the resultant PGM to calculate PGS in the UKBB^35,36^ data, to test its association with HGSOC and calculated its discriminatory ability for HGSOC by estimating the area under the ROC (AUROC) curve.

We assessed the performance of the PGS for women of East Asian ancestry using association summary statistics from BBJ. The reference panel consisted of individuals of East Asian ancestry from the 1000 Genomes Project^28^. This method has been described in detail elsewhere^53^. We also created candidate PGMs consisting of genotyped variants that could be more easily applied in clinical settings (Supplementary Methods).

### Absolute risks of EOC by PGS percentile

We calculated predicted lifetime risks (to age 80 years) of developing EOC for the general population, *BRCA1* and *BRCA2* PV carriers at the 1^st^, 5^th^, 20^th^, 50^th^ (median), 80^th^, 95^th^ and 99^th^ percentiles of the various PGS distributions, following previously published methodology^54^. To ensure consistency with known EOC risks for the general population, *BRCA1* and *BRCA2* PV carriers, average age-specific EOC incidence rates were constrained over PGS percentiles to agree with external EOC incidence rates for the general population^55^ and *BRCA1*/*2* carriers^56^.

We examined the number of *BRCA2* PV carriers genotyped on the OncoArray that would transition between risk groups (low (<10%) or high (≥10%) lifetime risk) of developing EOC, based on their observed PGS percentile. These risk reclassification analyses were limited to *BRCA2* carriers as their lifetime risks transition over the 10% lifetime risk threshold, whereas a *BRCA1* carrier is already at substantially increased lifetime risk.

### Ethics statement

All study participants provided written informed consent and participated in research or clinical studies at the host institute under ethically approved protocols. The studies and their approving institutes are listed in the Supplementary Material (Ethics Statement).

## RESULTS

The genome-wide association analyses for HGSOC were based on up to 398,238 women from OCAC (N=120,248, 30.2%), UKBB (N=245,377, 61.6%) and CIMBA (N=32,613, 8.2%) (Table 1, Supplementary Tables 1-2). A total of 19,883 (5.0%) women were classified as being diagnosed with HGSOC. The mean (standard deviation, SD) age at diagnosis for women in OCAC and UKBB were 60.2 (10.9) years and 63.5 (10.0) years, respectively. The mean (SD) censoring ages for BRCA1 and BRCA2 PV carriers were 43.7 (SD=12.0) years and 46.2 (SD=12.9) years, respectively.

### Reexamining previously identified associations with EOC

We looked up the associations for the lead variants previously identified as being associated with EOC in our new results (Supplementary Table 3, Supplementary Figure 1). Most lead variants previously reported to be associated specifically with HGSOC risk replicated in the present meta-analysis of OCAC, UKBB and *BRCA1/2* carriers at the significance threshold P<5×10^-8^. Exceptions were chr2:111138666 (rs17041869)^26^, chr2:113216387 (rs895412), chr11: 62126500 (rs7937840)^26^ and chr22:28538325 (rs6005807)^22^ (Supplementary Table 3). However, the chr2:110525257..111658369 and chr2:112716387..113716387 regions contained other variants that were associated at the genome-wide significance level in the present analysis, whilst the chr11:61626500..62626500 and chr22:28038325..29038325 regions did not contain any variants associated with HGSOC at the genome-wide significance level. It should be noted, however, that the chr2:111138666 (rs17041869) and chr11:62126500 (rs7937840) variants were identified through a cross-cancer (breast, ovarian, and prostate) GWAS^26^ and were not specifically identified as HGSOC associated variants.

### Novel loci associated with HGSOC

Associations with a total of 5,786 variants from 44 loci were significant at P<5×10^-8^. We excluded 5,778 variants at 37 loci from further consideration, as they were either near known associated regions (Supplementary Table 3), were not conditionally independent of the sentinel variant in a nearby known region, or were likely statistical artifacts arising from strata specific effects. There were eight variants associated (P<5×10^-8^) with HGSOC, at 5q11, 6p12, 8p21, 9p24-23, 16q22, 17p13 and 19q12 (Table 2, Figure 3, Supplementary Tables 4-5).

**Figure 3:**
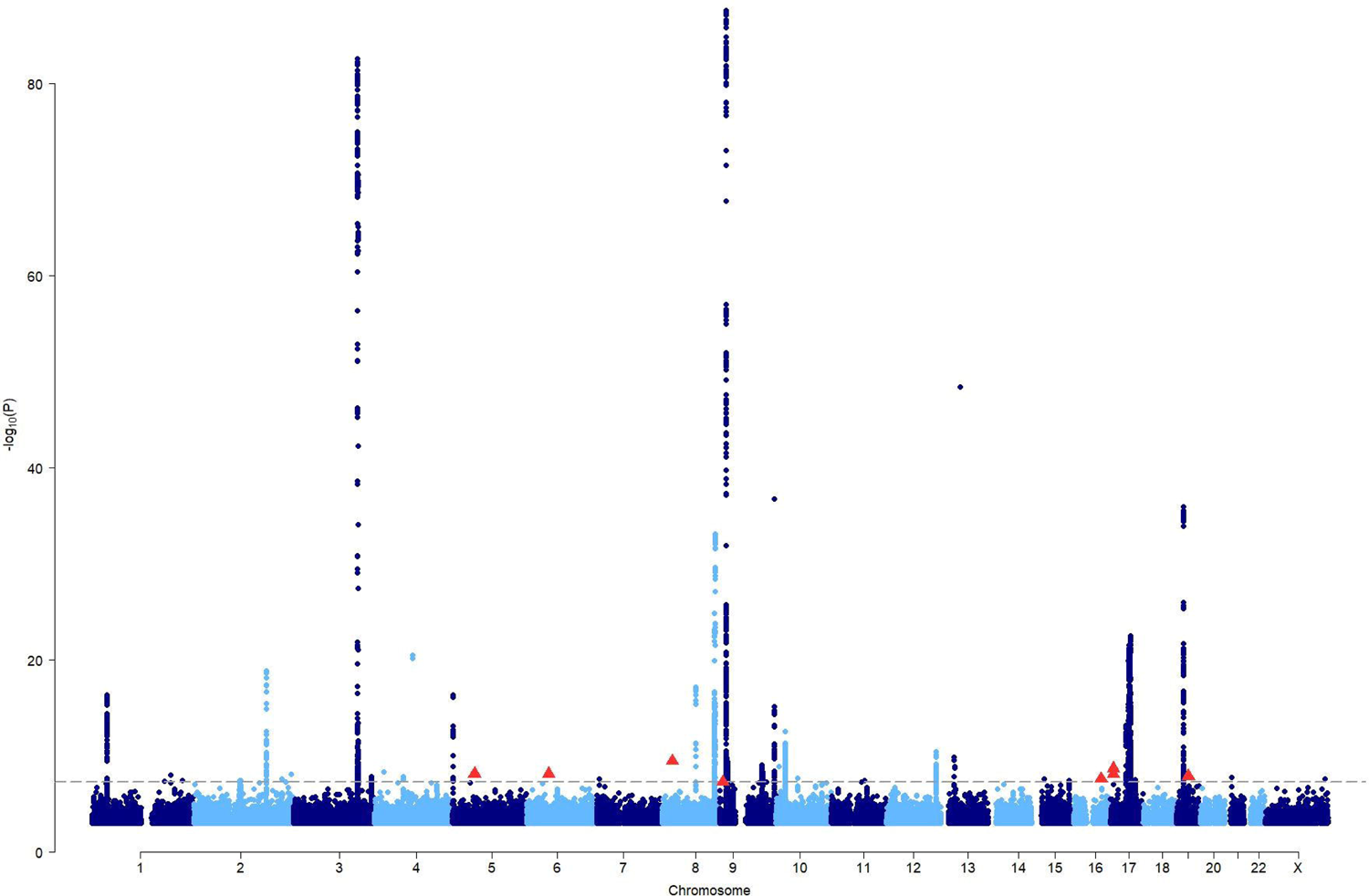
Manhattan plot showing the associations with HGSOC from the meta-analysis of OCAC, UKBB and CIMBA summary association data. The dashed line is the genome-wide statistical significance level (P=5×10^-8^). The eight independent genome-wide statistically significant variants at seven novel loci are shown as red triangles.

The associations at 5q11 (rs528577783-G; RR=5.15, 95%CI:2.96-8.96), 6p12 (rs1013698558-T; RR=2.35, 95%CI:1.76-3.14), 8p21 (rs191420549-A; RR=25.60, 95%CI:9.32-70.31), and 9p24-p23 (rs768719522-T; RR=10.04, 95%CI:4.38-22.99) were all single rare variants (MAF≤0.15%) associated with large HGSOC effects. The single SNPs associated with HGSOC at 16q22 (rs6979-G; RR=1.07, 95%CI:1.04-1.09) and 19q12 (rs62107113-A; RR=1.08, 95%CI:1.05-1.11) were common and conferred modest effects on HGSOC risk. There were two moderately correlated (TOPMed European^57^ *r*^2^=0.46, D’=0.89) low-frequency (MAF: 1.2% and 2.1%) variants at the 17p13 locus. The *TNFS13*/*TNFSF12*-*TNFSF13* intronic variant rs143094271-G was associated with a per-allele RR=1.28 (95%CI:1.18-1.39, P=7.61×10^-9^); and rs78378222-T, a *TP53* 3’-UTR variant, with a per-allele RR=1.44 (95%CI:1.28-1.62, P=1.76×10^-9^). The association effect size estimates at 16q22, 19q12 and 17p13 were consistent between OCAC/UKBB and *BRCA1/2* PV carriers. BFDPs indicated that the majority of these associations are likely to be true, although two rare variants, rs191420549 and rs768719522, had noticeably larger BFDPs (Table 2). Under a model assuming 1:1,000 truly associated variants, the BFDPs were 3.2% for rs191420549 and 11% for rs768719522. The other variants all had BFDP≤0.42%.

### Credible causal variants

We defined 52 CCVs across the seven novel regions (Supplementary Table 6, Supplementary Figure 2). Four regions (5q11, 6p12, 8p21, 9p24-23) had only the sentinel variant as a CCV, whilst the 16q22 (N=5), 17p13 (N=3) and 19q12 (N=40) loci had several CCVs.

### Association of the PGS with HGSOC

Of the 1,102 PGMs developed using OCAC and CIMBA data, the PGM that performed best in the FinnGen data comprised of 64,518 variants (Supplementary Table 7), and is denoted PGS_64518_. In the UKBB validation, the OR per SD of PGS_64518_ was estimated to be 1.46 (95%CI:1.37-1.54), with discriminatory ability of AUROC=0.607 (95%CI:0.590-0.623) (Table 3). The association of PGS_64518_ was strongly attenuated in the BBJ validation (East Asian ancestry women), where the OR per SD was 1.12 (95%CI:1.05-1.20).

When restricting the PGS to include only genotyped variants from the 64,518 genotyped and imputed variants, which may make their implementation easier, PGS with 5,957 (all genotyped variants from the 64,518) and 400 variants had similar performance characteristics. Relative to the PGS_64518_, a PGS considering the 400 most strongly associated genotyped variants, denoted PGS_400_, resulted in a small decrease in the AUROC to 0.603, and a marginally attenuated OR per SD (OR=1.43, 95%CI:1.35-1.52) in the UKBB.

### Predicted absolute risks for the general population and BRCA1/2 pathogenic variant carriers

The absolute lifetime risks (at age 80 years) for *BRCA1* PV carriers were predicted to be 25.9%, 42.8% and 64.7% at the 5^th^, 50^th^ and 95^th^ percentiles of the PGS_64518_ distribution, respectively (Table 3, Figure 4). The corresponding risks for PGS_64518_ for *BRCA2* PV carriers at the same PGS percentiles were predicted to be 9.3%, 16.7% and 28.9%, respectively (Table 3, Figure 4). The range of predicted percentile specific risks for the previously published 36 variant PGS^52^ was narrower, with risks for the same percentiles of 27.7%, 43.0% and 62.3% for *BRCA1* PV carriers, respectively, and 10.1%, 16.9% and 27.5% for *BRCA2* PV carriers, respectively. The PGS_400_ yielded absolute risks which were approximately at the midpoint of the 36 and 64,518 variant PGS absolute risks (*BRCA1* PV carriers: 26.7%, 42.9% and 63.6% at the 5^th^, 50^th^ and 95^th^ percentiles, respectively; *BRCA2* PV carriers: 9.7%, 16.8% and 28.3% at the 5^th^, 50^th^ and 95^th^ percentiles, respectively).

**Figure 4:**
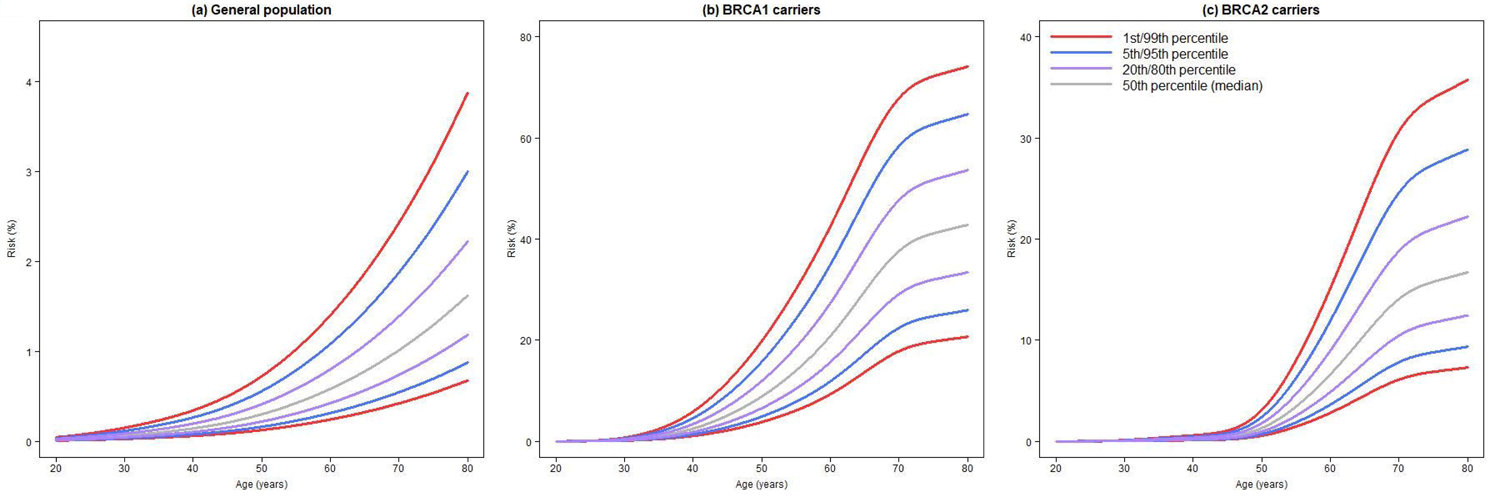
Predicted cumulative risks of developing EOC based on the PGS_64518_ at various percentiles of the PGS distribution for: (a) the general population (0.7% for 1^st^ percentile to 3.9% for the 99^th^ percentile), (b) *BRCA1* PV carriers (20.7% for 1^st^ percentile to 74.1% for the 99^th^ percentile), and (c) *BRCA2* PV carriers (7.3% for 1^st^ percentile to 35.7% for the 99^th^ percentile).

There was a net reclassification of *BRCA2* PV carriers at the 10% risk threshold when considering PGS_400_ and PGS_64518_ of 4.8% and 5.4%, respectively, compared to the 36 variant PGS (Supplementary Table 8).

## DISCUSSION

We conducted the largest GWAS to date for HGSOC, both in terms of the sample size and the number of genetic variants assessed. To do this, we made use of large international consortia (OCAC and CIMBA), and harnessed data from the UKBB to bolster the sample size. We also utilized recent advances in imputation reference panels, namely the TOPMed panel, which allowed us to investigate the largest number of genetic variants to date. We identified eight independent variants at seven loci to be genome-wide statistically significant for association with HGSOC risk, including four rare variants (MAF<1%) and two low-frequency polymorphisms (MAF 1-5%), demonstrating that GWAS with greater coverage imputation can identify previously undiscovered rare variant associations. Based on these associations, we defined 52 CCVs that have the strongest statistical evidence for being the likely causal variant for each locus. We also developed a polygenic model that exhibited improved discriminatory ability compared to previously published models.

The four rare variants were associated with large effect sizes, with RR estimates ranging from 2.35 to 25.6. The large effect sizes seem implausible and may reflect a winner’s curse effect with true effect sizes actually being lower than the estimates that we obtained^58^. Moreover, they may represent false positive associations, hence we estimated BFDPs^50^ for the eight novel variants to determine the likelihood of this. Six variants had low BFDPs, indicating they are likely true associations. However, 8p21 rs191420549 and 9p24-p23 rs768719522 had noticeably larger BFDPs and are more likely to represent false positive associations.

Variant rs78378222, in the *TP53* 3’-UTR, with the major T-allele (AF 98.8%) is associated with an increased risk of HGSOC. The same allele has been associated with an increased risk of triple-negative breast cancer^59^ with a per-allele relative risk of 1.45, similar to its association with HGSOC (RR=1.44). In contrast, the minor (G) allele of rs78378222 is associated with increased risks of skin^60–64^, brain^64–68^ and prostate cancers^60,69^. One study also found rs78378222 to be associated with standing height, lean body mass and basal metabolic rate^64^. The minor allele has been shown to impede *TP53* 3’-end processing, resulting in downregulated p53 mRNA levels and protein levels, and decreased apoptosis^70^. Furthermore, germline and somatic variations in *TP53* are well established factors in cancer development and progression through its role as a tumor suppressor^71–74^ and the *TP53* 3’-UTR germline variant has been shown to interact with tumor *TP53* mutation status^75^. A previous study by the OCAC found five SNPs in the *TP53* region, within ±21kb of rs78378222, to be associated with invasive EOC^76^. However, none of these SNPs are in LD with rs78378222.

rs528577783 is a rare intergenic variant between the *FST* and *NDUFS4* genes. *FST* encodes follistatin, a single-chain gonadal protein that specifically inhibits follicle-stimulating hormone release and is moderately expressed in human reproductive tissues (Supplementary Table 5)^77^. A linkage study identified *FST* as a candidate gene for polycystic ovary syndrome (PCOS)^78^. PCOS may be associated with an increased risk of borderline or postmenopausal ovarian cancer^79^, although a recent Mendelian Randomization study found that genetically predicted PCOS was associated with decreased risk of ovarian cancer^80^. A GWAS of African ancestry women found rs37792 near *FST* to be genome-wide statistically significantly associated with HGSOC in African ancestry women^81^.

The 6p12 variant rs1013698558, located ∼9.7kb from the *GCLC* gene, was moderately associated with HGSOC. A study found a synthetic lethality relationship between *GCLC* and *ARID1A*-deficient OC cells^82^, whilst another reports that *GCLC* inhibition causes apoptosis in *ARID1A*-deficient cancer cells^83^. *ARID1A* has been implicated as a tumor suppressor gene^84^. It may be that the association we find for rs1013698558 with HGSOC is a result of the interplay between *GCLC* and *ARID1A*. The 8p21 variant rs191420549 is 156kb from *CDCA2* and was associated with HGSOC with the largest RR that we report. *CDCA2* is involved in the cell division cycle and response to DNA damage. One study found *CDCA2* expression is upregulated in ovarian tumor tissue compared with normal tissue^85^. This study also found that *CDCA2* and its 100 most co-expressed genes were primarily involved in cell cycle, oocyte meiosis, progesterone-mediated oocyte maturation, p53 signaling and pyruvate metabolism pathways. We found that the *PTPRD* intronic variant rs768719522 at the 9p24-p23 locus had a large association RR with HGSOC. *PTPRD* has been implicated as a tumor suppressor gene^86^. This gene encodes a protein from the protein tyrosine phosphatase (PTP) family – PTPs are signaling molecules regulating processes such as cell growth, cell differentiation, the mitotic cycle and oncogenic transformation^87^. The common *ACD* missense variant rs6979 at 16q22 conferred a small per-allele RR to HGSOC risk. This gene encodes telomere protein TPP1, which is involved in maintenance of telomere length and protecting telomere ends. In addition to the 17p13 *TP53* 3’-UTR variant association, there was another independent variant associated with HGSOC risk at this locus. The rs143094271 variant is intronic in *TNFSF13*/*TNFSF12*-*TNFSF13*. These genes are members of the tumor necrosis factor family, which are involved in various cellular processes, including survival, proliferation, differentiation, and apoptosis. rs143094271-G has been associated with a decreased risk of having uterine fibroids (OR=0.70)^88^. Women with uterine fibroids have been found to be at increased risk of developing OC^89^. However, despite rs143094271-G being associated with women being less likely to have uterine fibroids, we found that rs143094271-G yielded an increased risk of developing HGSOC. The 19q12 common variant rs62107113 is located ∼15kb from the *CCNE1* gene and was associated with a modest increased risk of developing HGSOC. *CCNE1* encodes cyclin E1, which regulate cyclin-dependent kinases. Overexpression of *CCNE1* has been observed in genomically unstable tumors, notably HGSOC^90–94^ and triple-negative breast cancer^91,95–97^.

Moreover, it has been shown that replication stress in cells overexpressing *CCNE1* is likely a consequence of replication initiation, ultimately resulting in DNA damage and genomic instability^98^. There is evidence indicating *CCNE1* amplification is mutually exclusive to BRCA inactivation^92^. *CCNE1* is an exciting novel therapeutic target, Gallo *et al* developed an orally bioavailable PKMYT1 inhibitor that activated CDK1 in *CCNE1* overexpressed cells, promoting early mitosis in cells undergoing DNA synthesis^99^.

In addition to the 40 loci previously found to be associated with EOC^14–27^, we have identified genetic variants at a further seven loci associated with HGSOC, taking the number of loci associated with EOC to 47, 26 specifically with HGSOC. The previous known loci (52 variants at 40 loci) explained 8.5% of the polygenic variance of OC, assuming a total polygenic variance of 2.004 from Lee *et al*^100^. The six variants that we report to be associated with HGSOC with low BFDP explain an additional 2.8%, making the total variance explained by GWAS identified variants to be 11.3%.

For previously identified variants, the estimated associations from general population data (combined OCAC and UKBB) ORs were broadly consistent with the estimated HRs for *BRCA1* and *BRCA2* PV carriers (Supplementary Table 3). Furthermore, testing for heterogeneity of effects resulted in a small number of variants exhibiting differences in effect sizes between population-based ORs and carrier HRs. None of the eight novel associations exhibited any heterogeneous effects between population-based ORs and *BRCA1*/*2* PV carrier HRs (Supplementary Table 4).

Using the S4 method, we developed a 64,518 variant PGM, whose PGS was associated with a per SD OR of 1.46 (95%CI:1.37-1.54) and had discriminatory ability of AUROC=0.607 (95%CI:0.590-0.623). There was a small improvement over the previous best performing PGM developed by Dareng *et al*^52^ (18,007 variants; OR per unit SD = 1.42; AUROC=0.596) developed using similar methodology. The PGS_400_ could be more practical to implement in a clinical setting, since it contains fewer variants, all of which are known to successfully genotype from the OncoArray experiment, compared to the best performing PGS_64518_ which requires imputation.

We found that the PGS_400_ had similar performance to the optimal PGS_64518_. This also suggests that most of the predictive ability of the PGS may derive from genotyped SNPs with the largest variance contributions.

When each of the PGS were tested in individuals of East Asian ancestry from BBJ, the PGS associations were strongly attenuated, each to a similar degree. For women of East Asian ancestry, the PGS consisting of genotyped variants with 5,957 and 3,448 variants performed marginally better, with a slightly larger OR per PGS SD. The observed attenuations for East Asian women compared with European women are likely a result of the PGM derivation data being strongly weighted towards Europeans, as the majority of OCAC and CIMBA samples came from this ancestry group.

We calculated lifetime risks of developing EOC by PGS percentiles for the optimal 64,518 variant PGS for *BRCA1*/*2* carriers. The risks ranged from 25.9% to 64.7% for *BRCA1* carriers, and 9.3% to 28.9% for *BRCA2* carriers, at the 5^th^ and 95^th^ percentiles, respectively. The range of risks for the 36 variant PGS that is currently implemented in the CanRisk ovarian cancer risk prediction algorithm^52,100^ were narrower than those for the PGS_64518_. The lifetime risks based on the PGS_400_ that used a subset of 400 reliably genotyped variants included in the PGS_64518_ at these percentiles sat approximately at the midpoints of the risks from the 64,518 and 36 variant PGS. We compared what risk reclassification (lower risk, <10%, or higher risk, ≥10% lifetime risk) would occur when using the PGS_64518_ or PGS_400_ versus the 36 variant PGS for *BRCA2* PV carriers. We were unable to assess reclassification for *BRCA1* carriers as the lifetime absolute risks at the lowest percentiles of the PGS distributions were always above 10%. We found that the PGS_64518_ and PGS_400_ led to net reclassification of risk groups of around 5% versus the 36 variant PGS. Most reclassification shifted women from lower risk (<10%) using the 36 variant PGS to higher risk (≥10%) using the alternative PGS_64518_ or PGS_400_. Taken together, these estimated lifetime risks and risk reclassifications will be useful in more accurately determining a carriers’ risk and informing clinical management of risk.

Strengths of this study include using the TOPMed imputation reference panel, enabling us to assess a larger number of low-frequency and rare variants than previous studies. A further strength was its power to detect low-frequency and rare variant associations, as well as previously unidentified common variant associations. This was facilitated by additional genotyped samples included in both OCAC and CIMBA and using population-based data from UKBB, resulting in the largest sample size analysed for assessing genetic variant associations with HGSOC risk.

Limitations include the fact that the GWAS discovery data available were primarily of European ancestry; the associations of these variants are likely to differ for women of non-European ancestries, as they are likely to have different frequencies and LD patterns. A limitation of the PGM was that the derivation data differed from the GWAS discovery data presented here. Ideally, all the discovery GWAS data would have been used for PGM development. However, it was essential to validate the PGM on independent data, hence the OCAC and CIMBA data were used for development, whilst the UKBB data were reserved for validation. Lastly, the PGM training data (FinnGen) did not have specific histotypes available, meaning we were only able to consider overall EOC in the PGM training. As we were investigating non-mucinous OC, we would ideally have had specific EOC histotypes available at each stage of the PGM development, training and validation. However, given that HGSOC is the most prevalent EOC histotype, it is unlikely to have a major impact on the PGM hyperparameter fine-tuning.

Future research may aim to fine-map the novel loci identified here, to refine the candidate causal variants associated with HGSOC risk; and in-silico analyses may identify candidate target genes or pathways for further experimental studies^101^. Additionally, future research could aim to identify novel variants associated with other OC histotypes and to discover novel associations for other ancestries.

We have shown that improvements in imputation reference panels and increased sample sizes can identify novel HGSOC associated variants that previously went undetected, either from absence from genotyping or imputation reference panels, or from lack of power to detect associations. Furthermore, these associations can be used to develop PGM that outperform previous best efforts that can be incorporated into cancer risk prediction algorithms to improve personalized risk prediction for HGSOC.

## Data Availability

Summary statistics produced in the present study are available upon reasonable request to the authors.

## ACKNOWLEDGEMENTS AND FUNDING

This work was supported by Cancer Research UK grant: PPRPGM-Nov20\100002 and PRCPJT-May21\100006; by core funding from the NIHR Cambridge Biomedical Research Centre (NIHR203312) [*]. *The views expressed are those of the author(s) and not necessarily those of the NIHR or the Department of Health and Social Care.

## OCAC FUNDING

The Ovarian Cancer Association Consortium is supported by a grant from the Ovarian Cancer Research Fund thanks to donations by the family and friends of Kathryn Sladek Smith (PPD/RPCI.07). The scientific development and funding for this project were in part supported by the US National Cancer Institute GAME-ON Post-GWAS Initiative (U19-CA148112). This study made use of data generated by the Wellcome Trust Case Control consortium that was funded by the Wellcome Trust under award 076113.

The OCAC OncoArray genotyping project was funded through grants from the U.S. National Institutes of Health (CA1X01HG007491-01 (C.I.A.), U19-CA148112 (T.A.S.), R01-CA149429 (C.M.P.) and R01-CA058598 (M.T.G.); Canadian Institutes of Health Research (MOP-86727 (L.E.K.) and the Ovarian Cancer Research Fund (A.B.). The COGS project was funded through a European Commission’s Seventh Framework Programme grant (agreement number 223175 - HEALTH-F2-2009-223175).

Funding for individual studies: **AAS:** National Institutes of Health (RO1-CA142081); **AOV:** The Canadian Institutes for Health Research (MOP-86727); **AUS:** The Australian Ovarian Cancer Study Group was supported by the U.S. Army Medical Research and Materiel Command (DAMD17-01-1-0729), National Health & Medical Research Council of Australia (199600, 400413 and 400281), Cancer Councils of New South Wales, Victoria, Queensland, South Australia and Tasmania and Cancer Foundation of Western Australia (Multi-State Applications 191, 211 and 182). The Australian Ovarian Cancer Study gratefully acknowledges additional support from Ovarian Cancer Australia and the Peter MacCallum Foundation; **BAV:** ELAN Funds of the University of Erlangen-Nuremberg; **BEL:** National Kankerplan; **BGS:** Breast Cancer Now and the UK NIHR Biomedical Research Centre at the Institute of Cancer Research. **BVU:** National Institutes of Health/National Center for Advancing Translational Sciences (ULTR000445 and 1S10RR025141-01); **CAM:** National Institutes of Health Research Cambridge Biomedical Research Centre and Cancer Research UK Cambridge Cancer Centre; **CHA:** Innovative Research Team in University (PCSIRT) in China (IRT1076); **CNI:** Instituto de Salud Carlos III (PI 12/01319); Ministerio de Economía y Competitividad (SAF2012); **COE:** Department of Defense (W81XWH-11-2-0131); **CON:** National Institutes of Health (R01-CA063678, R01-CA074850; R01-CA080742); **DKE:** Ovarian Cancer Research Fund; **DOV:** National Institutes of Health (R01-CA112523 and R01-CA87538); **EMC:** Dutch Cancer Society (EMC 2014-6699); **EPC:** The coordination of EPIC is financially supported by the European Commission (DG-SANCO) and the International Agency for Research on Cancer. The national cohorts are supported by Danish Cancer Society (Denmark); Ligue Contre le Cancer, Institut Gustave Roussy, Mutuelle Générale de l’Education Nationale, Institut National de la Santé et de la Recherche Médicale (INSERM) (France); German Cancer Aid, German Cancer Research Center (DKFZ), Federal Ministry of Education and Research (BMBF) (Germany); the Hellenic Health Foundation (Greece); Associazione Italiana per la Ricerca sul Cancro-AIRC-Italy and National Research Council (Italy); Dutch Ministry of Public Health, Welfare and Sports (VWS), Netherlands Cancer Registry (NKR), LK Research Funds, Dutch Prevention Funds, Dutch ZON (Zorg Onderzoek Nederland), World Cancer Research Fund (WCRF), Statistics Netherlands (The Netherlands); ERC-2009-AdG 232997 and Nordforsk, Nordic Centre of Excellence programme on Food, Nutrition and Health (Norway); Health Research Fund (FIS), PI13/00061 to Granada, PI13/01162 to EPIC-Murcia, Regional Governments of Andalucía, Asturias, Basque Country, Murcia and Navarra, ISCIII RETIC (RD06/0020) (Spain); Swedish Cancer Society, Swedish Research Council and County Councils of Skåne and Västerbotten (Sweden); Cancer Research UK (14136, C570/A16491 and C8221/A19170), UK Medical Research Council (1000143 and MR/M012190/1); **GER:** German Federal Ministry of Education and Research, Programme of Clinical Biomedical Research (01 GB 9401) and the German Cancer Research Center; **GRC:** the European Union (European Social Fund - ESF) and Greek national funds through the Operational Program “Education and Lifelong Learning” of the National Strategic Reference Framework - Research Funding Program of the General Secretariat for Research & Technology (SYN11_10_19); **GRR:** Roswell Park Cancer Institute Alliance Foundation (P30 CA016056); **HAW:** U.S. National Institutes of Health (R01-CA58598, N01-CN-55424 and N01-PC-67001); **HJO:** German Research Foundation (Do 761/15-1); Rudolf-Bartling Foundation; **HMO:** Intramural funding; Rudolf-Bartling Foundation; **HOC:** Helsinki University Research Fund; **HOP:** Department of Defense (DAMD17-02-1-0669) and NCI (K07-CA080668, R01-CA95023, P50-CA159981 MO1-RR000056 R01-CA126841); **HUO:** German Research Foundation (Do 761/15-1); Rudolf-Bartling Foundation; **JGO:** JSPS KAKENHI grant; **JPN:** Grant-in-Aid for the Third Term Comprehensive 10-Year Strategy for Cancer Control from the Ministry of Health, Labour and Welfare; **KRA:** Korea Health Technology R&D Project through the Korea Health Industry Development Institute, and the National R&D Program for Cancer Control, Ministry of Health & Welfare, Republic of Korea (HI16C1127; 0920010); **LAX:** American Cancer Society Early Detection Professorship (SIOP-06-258-01-COUN) and the National Center for Advancing Translational Sciences (UL1TR000124); **LUN:** European Research Council (ERC-2011-AdG 294576), Swedish Cancer Society, Swedish Research Council, Beta Kamprad Foundation; **MAC:** National Institutes of Health (R01-CA122443, P30-CA15083, P50-CA136393); Mayo Foundation; Minnesota Ovarian Cancer Alliance; Fred C. and Katherine B. Andersen Foundation; Fraternal Order of Eagles; **MAL:** National Institutes for Health (R01-CA61107) Danish Cancer Society (94 222 52) from the, Copenhagen, Denmark; and the Mermaid I project; **MAS:** Malaysian Ministry of Higher Education (UM.C/HlR/MOHE/06) and Cancer Research Initiatives Foundation; **MAY:** National Institutes of Health (R01-CA122443, P30-CA15083, P50-CA136393); Mayo Foundation; Minnesota Ovarian Cancer Alliance; Fred C. and Katherine B. Andersen Foundation; **MCC:** Cancer Council Victoria, National Health and Medical Research Council of Australia (NHMRC) grants number 209057, 251533, 396414, and 504715; **MDA:** DOD Ovarian Cancer Research Program (W81XWH-07-0449); **MEC:** NIH (CA54281, CA164973, CA63464); **MOF:** Moffitt Cancer Center, Merck Pharmaceuticals, the state of Florida, Hillsborough County, and the city of Tampa; **NCO:** National Institutes of Health (R01-CA76016) and the Department of Defense (DAMD17-02-1-0666); **NEC:** National Institutes of Health R01-CA54419 and P50-CA105009 and Department of Defense W81XWH-10-1-02802; **NHS:** UM1 CA186107, P01 CA87969, R01 CA49449, R01-CA67262, UM1 CA176726; **NJO:** National Cancer Institute (NIH-K07 CA095666, R01-CA83918, NIH-K22-CA138563, and P30-CA072720) and the Cancer Institute of New Jersey; If Sara Olson and/or Irene Orlow is a co-author, please add NCI CCSG award (P30-CA008748) to the funding sources; **NOR:** Helse Vest, The Norwegian Cancer Society, The Research Council of Norway; **NTH:** Radboud University Medical Centre; **OPL:** National Health and Medical Research Council (NHMRC) of Australia (APP1025142) and Brisbane Women’s Club; **ORE:** OHSU Foundation; **OVA:** This work was supported by Canadian Institutes of Health Research grant (MOP-86727) and by NIH/NCI 1 R01CA160669-01A1; **PLC:** Intramural Research Program of the National Cancer Institute; **POC:** Pomeranian Medical University; **POL:** Intramural Research Program of the National Cancer Institute; **PVD:** Canadian Cancer Society and Cancer Research Society GRePEC Program; **RBH:** National Health and Medical Research Council of Australia; **RMH:** Cancer Research UK, Royal Marsden Hospital; **RPC:** National Institute of Health (P50 CA159981, R01CA126841); **SEA:** Cancer Research UK (C490/A10119 C490/A10124 C490/A16561); UK National Institute for Health Research Biomedical Research Centres at the University of Cambridge; **SIS:** NIH, National Institute of Environmental Health Sciences, Z01 ES044005 and Z01-ES049033; **SMC:** The Swedish Research Council; **SON:** National Health Research and Development Program, Health Canada, grant 6613-1415-53; **SRO:** Cancer Research UK (C536/A13086, C536/A6689) and Imperial Experimental Cancer Research Centre (C1312/A15589); **STA:** NIH grants U01 CA71966 and U01 CA69417; **SWE:** Swedish Cancer foundation, WeCanCureCancer and årKampMotCancer foundation; **SWH:** NIH (NCI) grant R37-CA070867; **TBO:** National Institutes of Health (R01-CA106414-A2), American Cancer Society (CRTG-00-196-01-CCE), Department of Defense (DAMD17-98-1-8659), Celma Mastery Ovarian Cancer Foundation; **TOR:** NIH grants R01 CA063678 and R01 CA063682; **UCI:** NIH R01-CA058860 and the Lon V Smith Foundation grant LVS-39420; **UHN:** Princess Margaret Cancer Centre Foundation-Bridge for the Cure; **UKO:** The UKOPS study was funded by The Eve Appeal (The Oak Foundation) and supported by the National Institute for Health Research University College London Hospitals Biomedical Research Centre; **UKR:** Cancer Research UK (C490/A6187), UK National Institute for Health Research Biomedical Research Centres at the University of Cambridge; **USC:** P01CA17054, P30CA14089, R01CA61132, N01PC67010, R03CA113148, R03CA115195, N01CN025403, and California Cancer Research Program (00-01389V-20170, 2II0200); **VAN:** BC Cancer Foundation, VGH & UBC Hospital Foundation; **VTL:** NIH K05-CA154337; **WMH:** National Health and Medical Research Council of Australia, Enabling Grants ID 310670 & ID 628903. Cancer Institute NSW Grants 12/RIG/1-17 & 15/RIG/1-16; **WOC:** National Science Centren (N N301 5645 40) The Maria Sklodowska-Curie Memorial Cancer Center and Institute of Oncology, Warsaw, Poland.

## OCAC ACKNOWLEDGEMENTS

We are grateful to the family and friends of Kathryn Sladek Smith for their generous support of the Ovarian Cancer Association Consortium through their donations to the Ovarian Cancer Research Fund. The OncoArray and COGS genotyping projects would not have been possible without the contributions of the following: Per Hall (COGS); Douglas F. Easton, Kyriaki Michailidou, Manjeet K. Bolla, Qin Wang (BCAC), Marjorie J. Riggan (OCAC), Rosalind A. Eeles, Ali Amin Al Olama, Zsofia Kote-Jarai, Sara Benlloch (PRACTICAL), Joe Dennis, Alison M. Dunning, Andrew Lee, Ed Dicks, Craig Luccarini and the staff of the Centre for Genetic Epidemiology Laboratory, Javier Benitez, Anna Gonzalez-Neira and the staff of the CNIO genotyping unit, Jacques Simard and Daniel C. Tessier, Francois Bacot, Daniel Vincent, Sylvie LaBoissière and Frederic Robidoux and the staff of the McGill University and Génome Québec Innovation Centre, Stig E. Bojesen, Sune F. Nielsen, Borge G. Nordestgaard, and the staff of the Copenhagen DNA laboratory, and Julie M. Cunningham, Sharon A. Windebank, Christopher A. Hilker, Jeffrey Meyer and the staff of Mayo Clinic Genotyping Core Facility. We thank the study participants, doctors, nurses, clinical and scientific collaborators, health care providers and health information sources who have contributed to the many studies contributing to this manuscript.

We thank all the women who have taken part in the contributing studies and we acknowledge the many study nurses, research assistants and all clinical and scientific collaborators. We thank the following individuals for their contribution to individual studies: **AOV:** Jennifer Koziak, Mie Konno, Michelle Darago, Faye Chambers and the Tom Baker Cancer Centre Translational Laboratories; **AUS:** The complete AOCS Study Group can be found at www.aocstudy.org **BEL:** Gilian Peuteman, Thomas Van Brussel, Annick Van den Broeck and Joke De Roover; **BVU:** The dataset(s) used for the analyses described were obtained from; **CAM:** This work was supported by Cancer Research UK; the University of Cambridge; National Institute for Health Research Cambridge Biomedical Research Centre; **CHA:** Innovative Research Team in University (PCSIRT) in China (IRT1076); **CHN:** To thank all members of Department of Obstetrics and Gynaecology, Hebei Medical University, Fourth Hospital and Department of Molecular Biology, Hebei Medical University, Fourth Hospital; **COE:** Gynecologic Cancer Center of Excellence (W81XWH-11-2-0131); **CON:** The cooperation of the 32 Connecticut hospitals, including Stamford Hospital, in allowing patient access, is gratefully acknowledged. This study was approved by the State of Connecticut Department of Public Health Human Investigation Committee. Certain data used in this study were obtained from the Connecticut Tumor Registry in the Connecticut Department of Public Health. The authors assume full responsibility for analyses and interpretation of these data; **DKE:** OCRF; **EPC:** To thank all members and investigators of the Rotterdam Ovarian Cancer Study. Dutch Cancer Society (EMC 2014-6699); **GER:** The German Ovarian Cancer Study (GER) thank Ursula Eilber for competent technical assistance; **HOC:** The study was supported by the Helsinki University Research Fund; **JGO:** JSPS KAKENHI grant; **KRA:** This study (Ko-EVE) was supported by a grant from the Korea Health Technology R&D Project through the Korea Health Industry Development Institute (KHIDI), and the National R&D Program for Cancer Control, Ministry of Health & Welfare, Republic of Korea (HI16C1127; 0920010); **LUN:** ERC - 2011-AdG, Swedish Cancer Society, Swedish Research Council; **MAS:** We would like to thank Famida Zulkifli and Ms Moey for assistance in patient recruitment, data collection and sample preparation. The Malaysian Ovarian Cancer Genetic Study is funded by research grants from the Malaysian Ministry of Higher Education (UM.C/HIR/MOHE/06) and charitable funding from Cancer Research Initiatives Foundation (MAY: P30-CA015083, P50-CA1363936, R01-CA248288); **MCC:** MCCS cohort recruitment was funded by VicHealth and Cancer Council Victoria. The MCCS was further supported by Australian NHMRC grants 209057, 251553 and 504711 and by infrastructure provided by Cancer Council Victoria. Cases and their vital status were ascertained through the Victorian Cancer Registry (VCR) and the Australian Institute of Health and Welfare (AIHW), including the National Death Index and the Australian Cancer Database; **MOF:** the Total Cancer Care™ Protocol and the Collaborative Data Services and Tissue Core Facilities at the H. Lee Moffitt Cancer Center & Research Institute, an NCI designated Comprehensive Cancer Center (P30-CA076292), Merck Pharmaceuticals and the state of Florida; **NHS:** The NHS/NHSII studies thank the following state cancer registries for their help: AL, AZ, AR, CA, CO, CT, DE, FL, GA, ID, IL, IN, IA, KY, LA, ME, MD, MA, MI, NE, NH, NJ, NY, NC, ND, OH, OK, OR, PA, RI, SC, TN, TX, VA, WA, and WY; **NJO: contact Elisa Bandera**; **OPL:** Members of the OPAL Study Group (http://opalstudy.qimrberghofer.edu.au/); **RPC:** National Institute of Health (P50 CA159981, R01CA126841); **SEA:** SEARCH team, Craig Luccarini, Caroline Baynes, Don Conroy; **SIS:** The Sister Study (SISTER) is supported by the Intramural Research Program of the NIH, National Institute of Environmental Health Sciences (Z01-ES044005 and Z01-ES049033); **SON:** National Health Research and Development Program, Health Canada, grant 6613-1415-53; **SRO:** To thank all members of Scottish Gynaecological Clinical Trails group and SCOTROC1 investigators; **SWE:** Swedish Cancer foundation, WeCanCureCancer and årKampMotCancer foundation; **SWH:** The SWHS is supported primarily by NIH grant R37-CA070867. We thank the participants and the research staff of the Shanghai Women’s Health Study for making this study possible; **UCI:** The UCI Ovarian cancer study is supported by the National Institutes of Health, National Cancer Institute grants CA58860, and the Lon V Smith Foundation grant LVS-39420; **UHN:** Princess Margaret Cancer Centre Foundation-Bridge for the Cure; **UKO:** We particularly thank I. Jacobs, M.Widschwendter, E. Wozniak, A. Ryan, J. Ford and N. Balogun for their contribution to the study**; UKR:** Carole Pye; **VAN:** BC Cancer Foundation, VGH & UBC Hospital Foundation; **WMH:** We thank the Gynaecological Oncology Biobank at Westmead, a member of the Australasian Biospecimen Network-Oncology group, which is funded by the National Health and Medical Research Council Enabling Grants ID 310670 & ID 628903 and the Cancer Institute NSW Grants 12/RIG/1-17 & 15/RIG/1-16 with financial support from the Sydney West Translational Cancer Research Centre, funded by the Cancer Institute NSW (15/TRC/1-01).

## CIMBA FUNDING

This work was supported by the Gray Foundation. GCT and ABS are NHMRC Research Fellows. iCOGS: the European Community’s Seventh Framework Programme under grant agreement n° 223175 (HEALTH-F2-2009-223175) (COGS), Cancer Research UK (C1287/A10118, C1287/A 10710, C12292/A11174, C1281/A12014, C5047/A8384, C5047/A15007, C5047/A10692, C8197/A16565), the National Institutes of Health (CA128978) and Post-Cancer GWAS initiative (1U19 CA148537, 1U19 CA148065 and 1U19 CA148112 - the GAME-ON initiative), the Department of Defence (W81XWH-10-1-0341), the Canadian Institutes of Health Research (CIHR) for the CIHR Team in Familial Risks of Breast Cancer (CRN-87521), and the Ministry of Economic Development, Innovation and Export Trade (PSR-SIIRI-701), Komen Foundation for the Cure, the Breast Cancer Research Foundation, and the Ovarian Cancer Research Fund. OncoArray: the PERSPECTIVE and PERSPECTIVE I&I projects funded by the Government of Canada through Genome Canada and the Canadian Institutes of Health Research, the ‘Ministère de l’Économie, de la Science et de l’Innovation du Québec’ through Genome Québec, and the Quebec Breast Cancer Foundation; the NCI Genetic Associations and Mechanisms in Oncology (GAME-ON) initiative and Discovery, Biology and Risk of Inherited Variants in Breast Cancer (DRIVE) project (NIH Grants U19 CA148065 and X01HG007492); and Cancer Research UK (C1287/A10118 and C1287/A16563).

ACBRCA: FAPESP/Fundação de Amparo à Pesquisa do Estado de São Paulo (2014/50943-1) São Paulo, Brazil, CNPq/Conselho Nacional de Desenvolvimento Científico e Tecnológico (465682/2014-6), Brazil. BCFR: U01 CA164920 from the National Cancer Institute. The content of this manuscript does not necessarily reflect the views or policies of the National Cancer Institute or any of the collaborating centers in the Breast Cancer Family Registry (BCFR), nor does mention of trade names, commercial products, or organizations imply endorsement by the US Government or the BCFR. BFBOCC: Lithuania (BFBOCC-LT): Research Council of Lithuania grant SEN-18/2015. BIDMC: Breast Cancer Research Foundation.

BMBSA: Cancer Association of South Africa (PI Elizabeth J. van Rensburg). BRICOH: SLN was partially supported by the Morris and Horowitz Families Endowed Professorship. CNIO: CNIO study is partially funded by Instituto de Salud Carlos III, project reference PI19/00640, cofunded by the European Regional Development Fund (ERDF), “A way to make Europe” and the Spanish Network on Rare Diseases (CIBERER). CCGCRN: Research reported in this publication was supported by the Breast Cancer Research Foundation (project 20-172), National Cancer Institute of the National Institutes of Health under grant number R25CA112486, and RC4CA153828 (PI: J. Weitzel) from the National Cancer Institute and the Office of the Director, National Institutes of Health. The content is solely the responsibility of the authors and does not necessarily represent the official views of the National Institutes of Health. CONSIT TEAM: Associazione Italiana Ricerca sul Cancro (AIRC; IG2015 no.16732) to P. Peterlongo. This work was partially supported by the Italian Ministry of Health with Ricerca Corrente and 5×1000 Funds (European Institute of Oncology). CZECANCA: Charles University/VFN projects Cooperatio - Laboratory Diagnostics/Medical Genetics, NU20-03-00285, NU20-09-00355, NU20-03-00016/ RVO-VFN 64165. DEMOKRITOS: European Union (European Social Fund – ESF) and Greek national funds through the Operational Program “Education and Lifelong Learning” of the National Strategic Reference Framework (NSRF) - Research Funding Program of the General Secretariat for Research & Technology: SYN11_10_19 NBCA. Investing in knowledge society through the European Social Fund. DFKZ: German Cancer Research Center. EMBRACE: Cancer Research UK Grants PRCPJT-Nov21\100004, C1287/A23382 and C1287/A26886. D. Gareth Evans and Fiona Lalloo are supported by an NIHR grant to the Biomedical Research Centre, Manchester. The Investigators at The Institute of Cancer Research and The Royal Marsden NHS Foundation Trust are supported by an NIHR grant to the Biomedical Research Centre at The Institute of Cancer Research and The Royal Marsden NHS Foundation Trust. Ros Eeles and Elizabeth Bancroft are supported by Cancer Research UK Grant C5047/A8385. Ros Eeles is also supported by NIHR support to the Biomedical Research Centre at The Institute of Cancer Research and The Royal Marsden NHS Foundation Trust. The FCCC and KUMC cohorts: The University of Kansas Cancer Center (P30 CA168524), the Kansas Institute for Precision Medicine (P20GM130423) and the Kansas Bioscience Authority Eminent Scholar Program. A.K.G. was funded by R01CA140323, R01CA214545, R01CA260132, 5U10CA180888, and by the Chancellors Distinguished Chair in Biomedical Sciences Professorship. FPGMX: A.Vega is supported by the Spanish Health Research Foundation, Instituto de Salud Carlos III (ISCIII), partially supported by FEDER funds through Research Activity Intensification Program (contract grant numbers: INT15/00070, INT16/00154, INT17/00133, INT20/00071), and through Centro de Investigación Biomédica en Red de Enferemdades Raras CIBERER (ACCI 2016: ER17P1AC7112/2018); Autonomous Government of Galicia (Consolidation and structuring program: IN607B), and by the Fundación Mutua Madrileña and Asociación Española Contra el Cáncer (AECC). GC-HBOC: German Cancer Aid (grant no 110837 and 113049, Rita K. Schmutzler), Federal Ministry of Education and Research, Germany (grant no 01GY1901), and the European Regional Development Fund and Free State of Saxony, Germany (LIFE - Leipzig Research Centre for Civilization Diseases, project numbers 713-241202, 713-241202, 14505/2470, 14575/2470). GEMO, a study from the National Cancer Genetics Network UNICANCER Genetic Group, France.: Ligue Nationale Contre le Cancer; the Association “Le cancer du sein, parlons-en!” Award, the Canadian Institutes of Health Research for the “CIHR Team in Familial Risks of Breast Cancer” program, the Fondation ARC pour la recherche sur le cancer (grant PJA 20151203365) and the French National Institute of Cancer (INCa grants AOR 01 082, 2001-2003, 2013-1-BCB-01-ICH-1 and SHS-E-SP 18-015). GEORGETOWN: the Non-Therapeutic Subject Registry Shared Resource at Georgetown University (NIH/NCI grant P30-CA051008), the Fisher Center for Hereditary Cancer and Clinical Genomics Research, and Swing Fore the Cure. HCSC: Spanish Ministry of Science and Innovation, ISCIII (Hayley) co-funded by FEDER Regional Development European Funds (EU). HEBCS: Helsinki University Hospital Research Fund, the Finnish Cancer Society and the Sigrid Juselius Foundation. The Hereditary Breast and Ovarian Cancer Research Group Netherlands (HEBON) consists of the following Collaborating Centers: Netherlands Cancer Institute; Erasmus Medical Center, Rotterdam; Leiden University Medical Center; Radboud University Nijmegen Medical Center; University Medical Center Utrecht; Amsterdam UMC, Univ of Amsterdam; Amsterdam UMC, Vrije Universiteit Amsterdam; Maastricht University Medical Center; University of Groningen; and acknowledges contributions of the Netherlands Comprehensive Cancer Organisation (IKNL), the nationwide network and registry of histo- and cytopathology in The Netherlands (PALGA), the Dutch Cancer Society grants NKI1998-1854, NKI2004-3088, NKI2007-3756, the Netherlands Organization of Scientific Research grant NWO 91109024, the Pink Ribbon grants 110005 and 2014-187.WO76, the BBMRI grant NWO 184.021.007/CP46 and the Transcan grant JTC 2012 Cancer 12-054. HEBCS: The Helsinki University Hospital Research Fund, The Sigrid Jusélius Foundation, The Cancer Foundation Finland. HRBCP: Hong Kong Sanatorium and Hospital, Dr Ellen Li Charitable Foundation, The Kerry Group Kuok Foundation, National Institute of Health1R 03CA130065, and North California Cancer Center. HUNBOCS: Hungarian Research Grants KTIA-OTKA CK-80745, NKFI_OTKA K-112228, and NKFI-FK-135065. HVH/VHIO: Orland Díez, Sara Gutiérrez-Enríquez and Judith Balmaña were funded by the Spanish Instituto de Salud Carlos III (ISCIII), an initiative of the Spanish Ministry of Economy and Innovation, partially supported by European Regional Development FEDER Funds, grant numbers PI16/01218, PI16/11363, PI19/01195 and PI19/01303 ICO: The authors would like to particularly acknowledge the support of the Asociación Española Contra el Cáncer (AECC), the Instituto de Salud Carlos III (organismo adscrito al Ministerio de Economía y Competitividad) and “Fondo Europeo de Desarrollo Regional (FEDER), una manera de hacer Europa” (PI10/01422, PI13/00285, PIE13/00022, PI15/00854, PI16/00563 and CIBERONC) and the Institut Català de la Salut and Autonomous Government of Catalonia (2009SGR290, 2014SGR338 and PERIS Project MedPerCan). ICARE: Inherited Cancer Registry (ICARE) based at the Vanderbilt-Ingram Cancer Center is funded, in part, through the Ingram Professorship. IFBCRC (Iranian Familial Breast Cancer Research Consortium) is funded by MCI (Motamed Cancer Institute) of ACECR. IHCC: PBZ_KBN_122/P05/2004 and the program of the Minister of Science and Higher Education under the name “Regional Initiative of Excellence” in 2019–2022 project number 002/RID /2018/19 amount of financing12 000 000 PLN.. INHERIT: Canadian Institutes of Health Research for the “CIHR Team in Familial Risks of Breast Cancer” program – grant # CRN-87521 and the Ministry of Economic Development, Innovation and Export Trade – grant # PSR-SIIRI-701. IOVHBOCS: Ministero della Salute and “5×1000” Istituto Oncologico Veneto grant. IPOBCS: Liga Portuguesa Contra o Cancro. kConFab: The National Breast Cancer Foundation, and previously by the National Health and Medical Research Council (NHMRC), the Queensland Cancer Fund, the Cancer Councils of New South Wales, Victoria, Tasmania and South Australia, and the Cancer Foundation of Western Australia. KOHBRA: the Korea Health Technology R&D Project through the Korea Health Industry Development Institute (KHIDI), and the National R&D Program for Cancer Control, Ministry of Health & Welfare, Republic of Korea (HI16C1127; 1020350; 1420190).

MAYO: NIH grants CA116167, CA192393, CA176785, and CA253187 an NCI Specialized Program of Research Excellence (SPORE) in Breast Cancer (CA116201),and a grant from the Breast Cancer Research Foundation. The MACBRCA study is funded by the Research Centre for Genetic Engineering and Biotechnology (RCGEB) “Georgi D. Efremov”, Macedonian Academy of Sciences and Arts (MASA). MCGILL: Jewish General Hospital Weekend to End Breast Cancer, Quebec Ministry of Economic Development, Innovation and Export Trade. Marc Tischkowitz is supported by an NIHR grant to the Biomedical Research Centre, Cambridge. MODSQUAD: MH CZ - DRO (MMCI, 00209805), and by Charles University in Prague project UNCE204024 (MZ). MSKCC: the Breast Cancer Research Foundation, the Robert and Kate Niehaus Clinical Cancer Genetics Initiative, the Andrew Sabin Research Fund and a Cancer Center Support Grant/Core Grant (P30 CA008748). NAROD: 1R01 CA149429-01. NCCS: NMRC Clinician Scientist Award (CSA), Terry Fox Foundation, Lee Foundation, PRECISE, Ministry of Education and NCCS Cancer Fund. NCI: the Intramural Research Program of the US National Cancer Institute, NIH, and by support services contracts NO2-CP-11019-50, N02-CP-21013-63 and N02-CP-65504 with Westat, Inc, Rockville, MD. NICCC: Clalit Health Services in Israel, the Israel Cancer Association and the Breast Cancer Research Foundation (BCRF), NY. NNPIO: Russian Science Foundation (grant 21-75-30015). NRG Oncology: U10 CA180868, NRG SDMC grant U10 CA180822, NRG Administrative Office and the NRG Tissue Bank (CA 27469), the NRG Statistical and Data Center (CA 37517) and the Intramural Research Program, NCI. OSUCCG: Ohio State University Comprehensive Cancer Center. OBRCA: funding from the Norwegian Cancer Society, contract 194751-2017. PBCS: Italian Association of Cancer Research (AIRC) [IG 2013 N.14477] and Tuscany Institute for Tumors (ITT) grant 2014-2015-2016. PCCM-CTCR is supported by the Corporate Social Responsibility Fund from Bajaj Auto Ltd, Grant GC-2528. SEABASS: Ministry of Science, Technology and Innovation, Ministry of Higher Education (UM.C/HlR/MOHE/06) and Cancer Research Initiatives Foundation. SGBCC was supported by the National Research Foundation Singapore (NRF-NRFF2017-02), NUS start-up Grant, National University Cancer Institute Singapore (NCIS) Centre Grant [NMRC/CG/NCIS/2010, NMRC/CG/012/2013, CGAug16M005], Breast Cancer Prevention Programme (BCPP), Asian Breast Cancer Research Fund, and the NMRC Clinician Scientist Award (SI Category) [NMRC/CSA-SI/0015/2017]. SMC: the Israeli Cancer Association. SWE-BRCA: the Swedish Cancer Society. UCHICAGO: NCI Specialized Program of Research Excellence (SPORE) in Breast Cancer (CA125183), R01 CA142996, 1U01CA161032 and by the Ralph and Marion Falk Medical Research Trust, the Entertainment Industry Fund National Women’s Cancer Research Alliance and the Breast Cancer research Foundation. OIO is an ACS Clinical Research Professor. UCLA: Jonsson Comprehensive Cancer Center Foundation; Breast Cancer Research Foundation. UCSF: UCSF Cancer Risk Program and Helen Diller Family Comprehensive Cancer Center. UKFOCR: Cancer Research UK. UPENN: Breast Cancer Research Foundation; Susan G. Komen Foundation for the cure, Basser Research Center for BRCA, NCI P30 CA016520.

UPITT/MWH: Hackers for Hope Pittsburgh. VFCTG: Victorian Cancer Agency, Cancer Australia, National Breast Cancer Foundation. WCP: Dr Karlan is funded by the American Cancer Society Early Detection Professorship (SIOP-06-258-01-COUN) and the National Center for Advancing Translational Sciences (NCATS), Grant UL1TR000124.

## CIMBA ACKNOWLEDGEMENTS

All the families and clinicians who contribute to the studies; Catherine M. Phelan for her contribution to CIMBA until she passed away on 22 September 2017; Sue Healey, in particular taking on the task of mutation classification with the late Olga Sinilnikova; clinicians, patients, researchers, technicians and nurses of A.C. Camargo Cancer Center for their contribution to this study; Oncogenetic Department, Clinical and Functional Genomics Group, Center of Genomic Diagnostics, Biobank and other International Research Center-CIPE’ facilities at AC. Camargo Cancer Center, especially Karina Miranda Santiago, Giovana Tardin Torrezan, José Claudio Casali, Nirvana Formiga and Fabiana Baroni Makdissi; Maggie Angelakos, Judi Maskiell, Gillian Dite, Helen Tsimiklis; members and participants in the New York site of the Breast Cancer Family Registry; members and participants in the Ontario Familial Breast Cancer Registry; Vilius Rudaitis and Laimonas Griškevičius; Drs Janis Eglitis, Anna Krilova and Aivars Stengrevics; Yuan Chun Ding and Linda Steele for their work in participant enrollment and biospecimen and data management; Bent Ejlertsen and Anne-Marie Gerdes for the recruitment and genetic counseling of participants; Alicia Barroso, Rosario Alonso and Guillermo Pita; all the individuals and the researchers who took part in CONSIT TEAM (Consorzio Italiano Tumori Ereditari Alla Mammella), in particular: Dario Zimbalatti, Daniela Zaffaroni, Laura Ottini, Giuseppe Giannini, Gabriele Lorenzo Capone, Liliana Varesco, Viviana Gismondi, Maria Grazia Tibiletti, Daniela Furlan, Antonella Savarese, Aline Martayan, Stefania Tommasi, Brunella Pilato, Bernardo Bonanni, Maria Rosaria Calvello, Irene Feroce, Monica Marabelli, Matilde Risti, Cristina Zanzottera, Loris Bernard, Elena Marino and the personnel of the Cogentech Cancer Genetic Test Laboratory, Milan, Italy. The FCCC cohort (Godwin) acknowledges Ms. JoEllen Weaver and Dr. Betsy Bove, and the KUMC cohort (Sharma and Godwin) acknowledge the support of Michele Park, Lauren DiMartino, Alex Webster and the current and past members of the Biospecimen Repository Core Facility (BRCF) at KUMC; all participants, clinicians, family doctors, researchers, and technicians for their contributions and commitment to the DKFZ study and the collaborating groups in Lahore, Pakistan (Noor Muhammad, Sidra Gull, Seerat Bajwa, Faiz Ali Khan, Humaira Naeemi, Saima Faisal, Asif Loya, Mohammed Aasim Yusuf) and Bogota, Colombia (Diana Torres, Ignacio Briceno, Fabian Gil). FPGMX: members of the Cancer Genetics group (IDIS): Ana Blanco, Miguel Aguado, Uxía Esperón and Belinda Rodríguez; the GIIS025 research nurses and staff for their contributions to this resource, and the many families who contribute to GIIS025; IFE - Leipzig Research Centre for Civilization Diseases (Markus Loeffler, Joachim Thiery, Matthias Nüchter, Ronny Baber); Genetic Modifiers of Cancer Risk in BRCA1/2 Mutation Carriers (GEMO) study is a study from the National Cancer Genetics Network UNICANCER Genetic Group, France. We wish to pay a tribute to Olga M. Sinilnikova, who with Dominique Stoppa-Lyonnet initiated and coordinated GEMO until she sadly passed away on the 30th June 2014. The team in Lyon (Olga Sinilnikova, Mélanie Léoné, Laure Barjhoux, Carole Verny-Pierre, Sylvie Mazoyer, Francesca Damiola, Valérie Sornin) managed the GEMO samples until the biological resource centre was transferred to Paris in December 2015 (Noura Mebirouk, Fabienne Lesueur, Dominique Stoppa-Lyonnet). We want to thank all the GEMO collaborating groups for their contribution to this study: Coordinating Centre, Service de Génétique, Institut Curie, Paris, France: Muriel Belotti, Ophélie Bertrand, Anne-Marie Birot, Bruno Buecher, Sandrine Caputo, Chrystelle Colas, Emmanuelle Fourme, Marion Gauthier-Villars, Lisa Golmard, Marine Le Mentec, Virginie Moncoutier, Antoine de Pauw, Claire Saule, Dominique Stoppa-Lyonnet, and Inserm U900, Institut Curie, Paris, France: Fabienne Lesueur, Noura Mebirouk, Yue Jiao.

Contributing Centres: Unité Mixte de Génétique Constitutionnelle des Cancers Fréquents, Hospices Civils de Lyon - Centre Léon Bérard, Lyon, France: Nadia Boutry-Kryza, Alain Calender, Sophie Giraud, Mélanie Léone. Institut Gustave Roussy, Villejuif, France: Brigitte Bressac-de-Paillerets, Odile Cabaret, Olivier Caron, Marine Guillaud-Bataille, Etienne Rouleau. Centre Jean Perrin, Clermont–Ferrand, France: Yves-Jean Bignon, Nancy Uhrhammer. Centre Léon Bérard, Lyon, France: Valérie Bonadona, Sophie Dussart, Christine Lasset, Pauline Rochefort. Centre François Baclesse, Caen, France: Pascaline Berthet, Laurent Castera, Dominique Vaur. Institut Paoli Calmettes, Marseille, France: Violaine Bourdon, Catherine Noguès, Tetsuro Noguchi, Cornel Popovici, Audrey Remenieras, Hagay Sobol. CHU Arnaud-de-Villeneuve, Montpellier, France: Isabelle Coupier, Pascal Pujol. Centre Oscar Lambret, Lille, France: Claude Adenis, Aurélie Dumont, Françoise Révillion.

Centre Paul Strauss, Strasbourg, France: Danièle Muller. Institut Bergonié, Bordeaux, France: Emmanuelle Barouk-Simonet, Françoise Bonnet, Virginie Bubien, Anaïs Dupré, Anne Floquet, Michel Longy, Marie Louty, Cécile Maninna, Nicolas Sevenet, Institut Claudius Regaud, Toulouse, France: Laurence Gladieff, Rosine Guimbaud, Viviane Feillel, Christine Toulas. CHU Grenoble, France: Hélène Dreyfus, Dominique Leroux, Clémentine Legrand, Christine Rebischung. CHU Dijon, France: Amandine Baurand, Geoffrey Bertolone, Fanny Coron, Laurence Faivre, Caroline Jacquot, Sarab Lizard, Sophie Nambot. CHU St-Etienne, France: Caroline Kientz, Marine Lebrun, Fabienne Prieur. Hôtel Dieu Centre Hospitalier, Chambéry, France: Sandra Fert Ferrer. Centre Antoine Lacassagne, Nice, France: Véronique Mari. CHU Limoges, France: Laurence Vénat-Bouvet. CHU Nantes, France: Stéphane Bézieau, Capucine Delnatte. CHU Bretonneau, Tours and Centre Hospitalier de Bourges France: Isabelle Mortemousque. Groupe Hospitalier Pitié-Salpétrière, Paris, France: Florence Coulet, Mathilde Warcoin. CHU Vandoeuvre-les-Nancy, France: Myriam Bronner, Johanna Sokolowska. CHU Besançon, France: Marie-Agnès Collonge-Rame. CHU Poitiers, Centre Hospitalier d’Angoulême and Centre Hospitalier de Niort, France: Stéphanie Chieze-Valero, Paul Gesta, Brigitte Gilbert-Dussardier. Centre Hospitalier de La Rochelle: Hakima Lallaoui. CHU Nîmes Carémeau, France: Jean Chiesa. CHI Poissy, France: Denise Molina-Gomes. CHU Angers, France: Olivier Ingster; CHU de Martinique, France: Odile Bera; Mickaelle Rose; Drs. Taru A. Muranen and Carl Blomqvist, RN Outi Malkavaara;; The Hereditary Breast and Ovarian Cancer Research Group Netherlands (HEBON) consists of the following Collaborating Centers: Netherlands Cancer Institute (coordinating center), Amsterdam, NL: M.A. Rookus, F.B.L. Hogervorst, F.E. van Leeuwen, M.A. Adank, M.K. Schmidt, D.J. Jenner; Erasmus Medical Center, Rotterdam, NL: J.M. Collée, A.M.W. van den Ouweland, M.J. Hooning, I.A. Boere; Leiden University Medical Center, NL: C.J. van Asperen, P. Devilee, R.B. van der Luijt, T.C.T.E.F. van Cronenburg; Radboud University Nijmegen Medical Center, NL: M.R. Wevers, A.R. Mensenkamp; University Medical Center Utrecht, NL: M.G.E.M. Ausems, M.J. Koudijs; Amsterdam UMC, Univ of Amsterdam, NL: I. van de Beek; Amsterdam UMC, Vrije Universiteit Amsterdam, NL: K. van Engelen, J.J.P. Gille; Maastricht University Medical Center, NL: E.B. Gómez García, M.J. Blok, M. de Boer; University of Groningen, NL: L.P.V. Berger, A.H. van der Hout, M.J.E. Mourits, G.H. de Bock; The Netherlands Comprehensive Cancer Organisation (IKNL): S. Siesling, J. Verloop; The nationwide network and registry of histo- and cytopathology in The Netherlands (PALGA): E.C. van den Broek; the study participants and the registration teams of IKNL and PALGA for part of the HEBON data collection; Hong Kong Sanatorium and Hospital; the Hungarian Breast and Ovarian Cancer Study Group members (Attila Patócs, János Papp, Anikó Bozsik, Timea Pócza, Zoltán Mátrai, Lajos Géczi, National Institute of Oncology, Budapest, Hungary) and the clinicians and patients for their contributions to this study; Fatemeh Yadegari, Shiva Zarinfam and Rezvan Esmaeili for their role in participant enrollment and biospecimen and data management; the study participants and registration teams of the Hereditary Cancer Genetics Group of the Valld’Hebron Institute of Oncolgy (VHIO) and the Clinical and Molecular Genetics Department of the University Hospital Vall d’Hebron (HVH), the Cellex Foundation for providing research facilities, and CERCA Programme/Generalitat de Catalunya for institutional support; members and participants of the Inherited Cancer Registry (ICARE); the ICO Hereditary Cancer Program team led by Dr. Gabriel Capella; the ICO Hereditary Cancer Program team led by Dr. Gabriel Capella; Dr Martine Dumont for sample management and skillful assistance; Catarina Santos and Pedro Pinto; members of the Center of Molecular Diagnosis, Oncogenetics Department and Molecular Oncology Research Center of Barretos Cancer Hospital; Heather Thorne, Eveline Niedermayr, all the kConFab research nurses and staff, the heads and staff of the Family Cancer Clinics, and the Clinical Follow Up Study (which has received funding from the NHMRC, the National Breast Cancer Foundation, Cancer Australia, and the National Institute of Health (USA)) for their contributions to this resource, and the many families who contribute to kConFab; the KOBRA Study Group; all participants and the collaborators from RCGEB “Georgi D. Efremov”, MASA (Ivana Maleva Kostovska, Simona Jakovcevska, Sanja Kiprijanovska), University Clinic of Radiotherapy and Oncology (Snezhana Smichkoska, Emilija Lazarova, Marina Iljovska), Adzibadem-Sistina Hospital (Katerina Kubelka-Sabit, Dzengis Jasar, Mitko Karadjozov), and Re-Medika Hospital (Andrej Arsovski and Liljana Stojanovska) for their contributions and commitment to the MACBRCA study; Csilla Szabo (National Human Genome Research Institute, National Institutes of Health, Bethesda, MD, USA); Eva Machackova (Department of Cancer Epidemiology and Genetics, Masaryk Memorial Cancer Institute and MF MU, Brno, Czech Republic); Petra Kleiblova, Marketa Janatova, Jana Soukupova (Institute of Medical Biochemistry and Laboratory Diagnostics, 1st Faculty of Medicine, Charles University and General University Hospital in Prague (VFN), Czechia), Petra Zemankova, Petr Nehasil (Institute of Pathological Physiology, 1st Faculty of Medicine, Charles University, Czechia), Michal Vocka (Department of Oncology, General University Hospital in Prague (VFN), Czechia), Anne Lincoln, Lauren Jacobs; the participants in Hereditary Breast/Ovarian Cancer Study and Breast Imaging Study for their selfless contributions to our research; the NICCC National Familial Cancer Consultation Service team led by Sara Dishon, the lab team led by Dr. Flavio Lejbkowicz, and the research field operations team led by Dr. Mila Pinchev; the staff of Genetic Health Service NZ and the families who have contributed; members and participants in the Ontario Cancer Genetics Network; Hayley Cassingham. Leigha Senter, Kevin Sweet, Julia Cooper, and Amber Aielts; research nurses and staff of Breast Unit, Pauls Stradins Clinical University Hopsital, RSUIO and the many families who contribute to the CIMBA registry of RSUIO; Yip Cheng Har, Nur Aishah Mohd Taib, Phuah Sze Yee, Norhashimah Hassan and all the research nurses, research assistants and doctors involved in the MyBrCa Study for assistance in patient recruitment, data collection and sample preparation, Philip Iau, Sng Jen-Hwei and Sharifah Nor Akmal for contributing samples from the Singapore Breast Cancer Study and the HUKM-HKL Study respectively; the National Cancer Centre Singapore Cancer Genetics Service (NCCS) for patient recruitement; the Meirav Comprehensive breast cancer center team at the Sheba Medical Center; Christina Selkirk; Åke Borg, Håkan Olsson, Helena Jernström, Karin Henriksson, Katja Harbst, Maria Soller, Ulf Kristoffersson; from Gothenburg Sahlgrenska University Hospital: Anna Öfverholm, Margareta Nordling, Per Karlsson, Zakaria Einbeigi; from Stockholm and Karolinska University Hospital: Anna von Wachenfeldt, Annelie Liljegren, Annika Lindblom, Brita Arver, Gisela Barbany Bustinza, Johanna Rantala; from Umeå University Hospital: Beatrice Melin, Christina Edwinsdotter Ardnor, Monica Emanuelsson; from Uppsala University: Hans Ehrencrona, Maritta Hellström Pigg, Richard Rosenquist; from Linköping University Hospital: Marie Stenmark-Askmalm, Sigrun Liedgren; Cecilia Zvocec, Qun Niu; Joyce Seldon and Lorna Kwan; Dr. Robert Nussbaum, Beth Crawford, Kate Loranger, Julie Mak, Nicola Stewart, Robin Lee, Amie Blanco and Peggy Conrad and Salina Chan; Patricia Harrington; Geoffrey Lindeman, Marion Harris, Joanne McKinley, Simone McInerny, and Ella Thompson for performing all DNA amplification.

This research has been conducted using the UK Biobank Resource under Application Number 28126. We want to acknowledge the participants and investigators of the FinnGen study. The BioBank Japan Project was supported by the Tailor-Made Medical Treatment program of the Ministry of Education, Culture, Sports, Science, and Technology (MEXT), the Japan Agency for Medical Research and Development (AMED). The Genotype-Tissue Expression (GTEx) Project was supported by the Common Fund of the Office of the Director of the National Institutes of Health, and by NCI, NHGRI, NHLBI, NIDA, NIMH, and NINDS. The data used for the analyses described in this manuscript were obtained from: https://gtexportal.org/home/gene/ the GTEx Portal on 03/31/2023.

## SUPPLEMENTARY TABLE LEGENDS

*Supplementary Table 1*: Numbers of controls and EOC cases by genotyping array and country from population-based studies. The “cases” column (column E) is a total of the histotypes in columns F-K. LMP = low malignant potential, HGSOC = high-grade serous ovarian cancer, ENOC = endometrioid ovarian cancer, CCOC = clear cell ovarian cancer, MOC = mucinous ovarian cancer.

*Supplementary Table 2*: Numbers of unaffected *BRCA1*/*2* carriers and carriers diagnosed with EOC, by CIMBA study.

*Supplementary Table 3*: Previously identified genomic regions associated with EOC. Lookups are presented for the variant’s association with its most strongly associated EOC histotype from the OCAC. Lookups are also presented from the present HGSOC meta-analysis of OCAC, UKBB and *BRCA1*/*2* carriers, and its component parts (associations from OCAC and UKBB combined, associations from *BRCA1* carriers, and associations from *BRCA2* carriers). Chr = chromosome. Positions are on build 38. SNP = rsID of the most strongly associated variant with “best histotype”. Alleles = reference/effect alleles. EAF = effect allele frequency. RR = relative risk (per effect allele). CI = confidence interval. P = P-value for association with HGSOC. P_het_ = P-value for heterogeneity between the general population OR (OCAC and UKBB meta-analysis, column U) and the PV carrier HRs (*BRCA1* PV carriers, column Z; *BRCA2* PV carriers, column AF). SNPs and positions (columns E-F) highlighted in yellow are most strongly associated with HGSOC in OCAC that did not replicate (P<5×10^-8^) in the OCAC+UKBB+CIMBA meta-analysis (columns N and S). Histotypes: NMOC = non-mucinous, HGSOC = high-grade serous, LGSOC = low-grade serous, MOC = mucinous, CCOC = clear cell, EnOC = endometrioid. * Previously identified through meta-analysis of OCAC and CIMBA. ** Previously identified through multi-cancer meta-analysis. *** When combining result with most recent results from CIMBA (unpublished).

*Supplementary Table 4*: Novel associations by major analysis units (OCAC, UKBB, *BRCA1* carriers and *BRCA2* carriers). Positions are on build 38. EAF = effect allele frequency. OR = odds ratio per effect allele. HR = hazard ratio per effect allele. CI = confidence interval. P = P-value for association. UKBB did not contribute associations for rs1013698558 (chr6:53554782:A:T). * HRs were estimated but had large estimated standard errors resulting in wide CIs – these associations were used in the meta-analysis between OCAC, UKBB and CIMBA, but contributed little as the inverse variance weights were extremely small.

*Supplementary Table 5*: Further details for eight novel variants associated with HGSOC risk. Positions are on build 38. Gene descriptions are taken from dbSNP^87^. Phenoscanner^102–104^ and PheWeb^105^ are lookups of relevant associations for these variants/genes. GTEx^77^ gene expression data is reported for human reproductive tissues with TPM (transcripts per million) > 5. eQTLGen^106,107^ lookups for cis- and trans-eQTLs for variants and genes were performed.

*Supplementary Table 6*: List of credible causal variants (CCVs) at each novel region. Sentinel variants are highlighted in green. These data are presented graphically in Supplementary Figure 2.

*Supplementary Table 7*: List of variants contributing to the polygenic models (PGMs). Positions are on build 38. Weights are given for five PGMs and are per effect allele. Instances in which weights are not present (NA) means that that variant does not contribute to that PGM. The weights are taken from the OCAC, UKBB and CIMBA meta-analysis effect size estimates with the hyperparameters applied. Four of the eight variants found in the discovery GWAS were directly included in the 64,518 variant PGM (rs6979 chr16:67657765, rs143094271 chr17:7559785, rs78378222 chr17:7668434 and rs62107113 chr19:29797136, highlighted in green in the table).

*Supplementary Table 8*: Risk reclassification based on different polygenic scores for *BRCA2* carriers. Lifetime risks were categorized as lower risk (lifetime risk below 10%) or higher risk (10% or greater). The PGM_64518_ and PGM_400_ were compared with the 36 SNP PGS currently used in the CanRisk prediction algorithm.

## SUPPLEMENTARY FIGURE LEGENDS

**Supplementary Figure 1:**
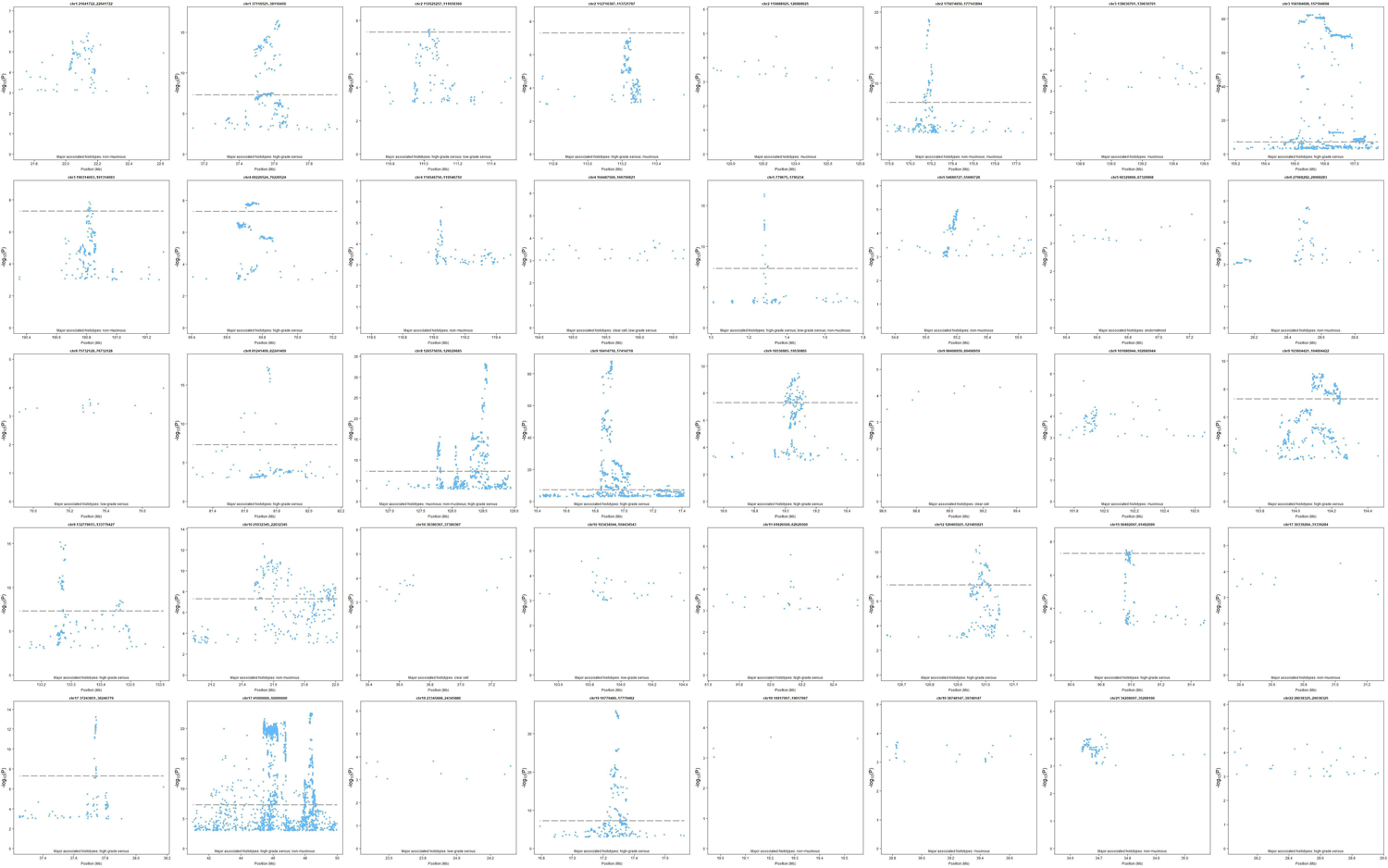
Regional association plots for 40 genomic regions previously associated with any EOC histotype. Regions are described in Supplementary Table 3.

**Supplementary Figure 2:**
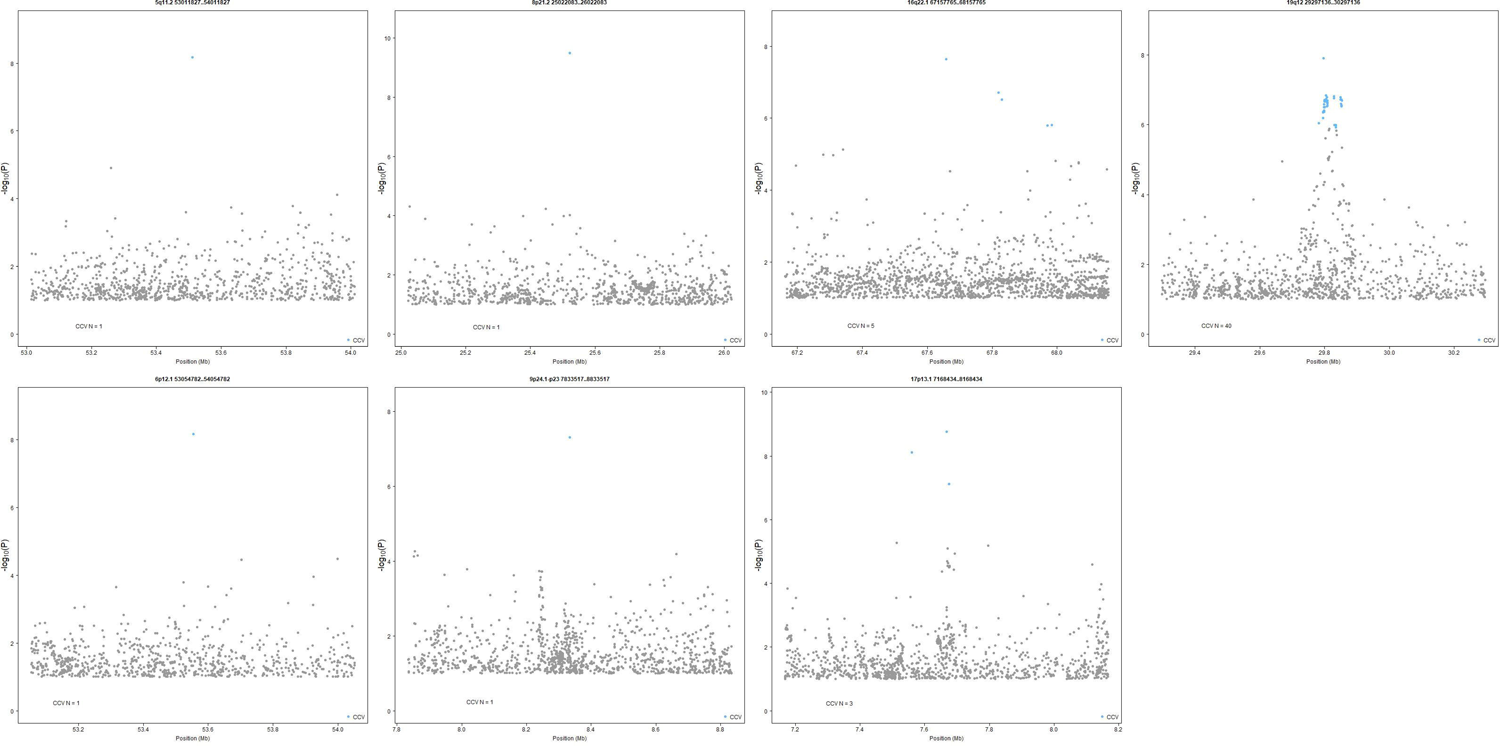
Regional association plots for seven novel genomic regions associated with HGSOC. Credible causal variants (CCVs) are highlighted in blue. Lists of CCVs are presented in Supplementary Table 6.

## REFERENCES

1. Sung H, Ferlay J, Siegel RL, et al. Global Cancer Statistics 2020: GLOBOCAN Estimates of Incidence and Mortality Worldwide for 36 Cancers in 185 Countries. CA Cancer J Clin 2021;71(3):209–249. DOI: 10.3322/caac.21660.

2. Prat J. New insights into ovarian cancer pathology. Ann Oncol 2012;23 Suppl 10:x111–7. DOI: 10.1093/annonc/mds300.

3. Peres LC, Cushing-Haugen KL, Kobel M, et al. Invasive Epithelial Ovarian Cancer Survival by Histotype and Disease Stage. J Natl Cancer Inst 2019;111(1):60–68. DOI: 10.1093/jnci/djy071.

4. Risch HA, McLaughlin JR, Cole DE, et al. Population BRCA1 and BRCA2 mutation frequencies and cancer penetrances: a kin-cohort study in Ontario, Canada. J Natl Cancer Inst 2006;98(23):1694–706. DOI: 10.1093/jnci/djj465.

5. Risch HA, McLaughlin JR, Cole DE, et al. Prevalence and penetrance of germline BRCA1 and BRCA2 mutations in a population series of 649 women with ovarian cancer. Am J Hum Genet 2001;68(3):700–10. DOI: 10.1086/318787.

6. Shaw PA, McLaughlin JR, Zweemer RP, et al. Histopathologic features of genetically determined ovarian cancer. Int J Gynecol Pathol 2002;21(4):407–11. DOI: 10.1097/00004347-200210000-00011.

7. Zhang S, Royer R, Li S, et al. Frequencies of BRCA1 and BRCA2 mutations among 1,342 unselected patients with invasive ovarian cancer. Gynecol Oncol 2011;121(2):353–7. DOI: 10.1016/j.ygyno.2011.01.020.

8. Schrader KA, Hurlburt J, Kalloger SE, et al. Germline BRCA1 and BRCA2 mutations in ovarian cancer: utility of a histology-based referral strategy. Obstet Gynecol 2012;120(2 Pt 1):235–40. DOI: 10.1097/AOG.0b013e31825f3576.

9. Wang YK, Bashashati A, Anglesio MS, et al. Genomic consequences of aberrant DNA repair mechanisms stratify ovarian cancer histotypes. Nat Genet 2017;49(6):856–865. DOI: 10.1038/ng.3849.

10. Lakhani SR, Manek S, Penault-Llorca F, et al. Pathology of ovarian cancers in BRCA1 and BRCA2 carriers. Clin Cancer Res 2004;10(7):2473–81. DOI: 10.1158/1078-0432.ccr-1029-3.

11. O’Mahony DG, Ramus SJ, Southey MC, et al. Ovarian cancer pathology characteristics as predictors of variant pathogenicity in BRCA1 and BRCA2. Br J Cancer 2023;128(12):2283–2294. DOI: 10.1038/s41416-023-02263-5.

12. Singh N, McCluggage WG, Gilks CB. High-grade serous carcinoma of tubo-ovarian origin: recent developments. Histopathology 2017;71(3):339–356. DOI: 10.1111/his.13248.

13. Kobel M, Kalloger SE, Huntsman DG, et al. Differences in tumor type in low-stage versus high-stage ovarian carcinomas. Int J Gynecol Pathol 2010;29(3):203–11. DOI: 10.1097/PGP.0b013e3181c042b6.

14. Song H, Ramus SJ, Tyrer J, et al. A genome-wide association study identifies a new ovarian cancer susceptibility locus on 9p22.2. Nat Genet 2009;41(9):996–1000. DOI: 10.1038/ng.424.

15. Bolton KL, Tyrer J, Song H, et al. Common variants at 19p13 are associated with susceptibility to ovarian cancer. Nat Genet 2010;42(10):880–4. DOI: 10.1038/ng.666.

16. Goode EL, Chenevix-Trench G, Song H, et al. A genome-wide association study identifies susceptibility loci for ovarian cancer at 2q31 and 8q24. Nat Genet 2010;42(10):874–9. DOI: 10.1038/ng.668.

17. Bojesen SE, Pooley KA, Johnatty SE, et al. Multiple independent variants at the TERT locus are associated with telomere length and risks of breast and ovarian cancer. Nat Genet 2013;45(4):371–84, 384e1-2. DOI: 10.1038/ng.2566.

18. Permuth-Wey J, Lawrenson K, Shen HC, et al. Identification and molecular characterization of a new ovarian cancer susceptibility locus at 17q21.31. Nat Commun 2013;4:1627. DOI: 10.1038/ncomms2613.

19. Pharoah PD, Tsai YY, Ramus SJ, et al. GWAS meta-analysis and replication identifies three new susceptibility loci for ovarian cancer. Nat Genet 2013;45(4):362–70, 370e1-2. DOI: 10.1038/ng.2564.

20. Shen H, Fridley BL, Song H, et al. Epigenetic analysis leads to identification of HNF1B as a subtype-specific susceptibility gene for ovarian cancer. Nat Commun 2013;4:1628. DOI: 10.1038/ncomms2629.

21. Kuchenbaecker KB, Ramus SJ, Tyrer J, et al. Identification of six new susceptibility loci for invasive epithelial ovarian cancer. Nat Genet 2015;47(2):164–71. DOI: 10.1038/ng.3185.

22. Phelan CM, Kuchenbaecker KB, Tyrer JP, et al. Identification of 12 new susceptibility loci for different histotypes of epithelial ovarian cancer. Nat Genet 2017;49(5):680–691. DOI: 10.1038/ng.3826.

23. Couch FJ, Wang X, McGuffog L, et al. Genome-wide association study in BRCA1 mutation carriers identifies novel loci associated with breast and ovarian cancer risk. PLoS Genet 2013;9(3):e1003212. DOI: 10.1371/journal.pgen.1003212.

24. Chen K, Ma H, Li L, et al. Genome-wide association study identifies new susceptibility loci for epithelial ovarian cancer in Han Chinese women. Nat Commun 2014;5:4682. DOI: 10.1038/ncomms5682.

25. Kelemen LE, Lawrenson K, Tyrer J, et al. Genome-wide significant risk associations for mucinous ovarian carcinoma. Nat Genet 2015;47(8):888–97. DOI: 10.1038/ng.3336.

26. Kar SP, Beesley J, Amin Al Olama A, et al. Genome-Wide Meta-Analyses of Breast, Ovarian, and Prostate Cancer Association Studies Identify Multiple New Susceptibility Loci Shared by at Least Two Cancer Types. Cancer Discov 2016;6(9):1052–67. DOI: 10.1158/2159-8290.CD-15-1227.

27. Lawrenson K, Kar S, McCue K, et al. Functional mechanisms underlying pleiotropic risk alleles at the 19p13.1 breast-ovarian cancer susceptibility locus. Nat Commun 2016;7:12675. DOI: 10.1038/ncomms12675.

28. Genomes Project Consortium, Auton A, Brooks LD, et al. A global reference for human genetic variation. Nature 2015;526(7571):68–74. DOI: 10.1038/nature15393.

29. McCarthy S, Das S, Kretzschmar W, et al. A reference panel of 64,976 haplotypes for genotype imputation. Nat Genet 2016;48(10):1279–83. DOI: 10.1038/ng.3643.

30. Taliun D, Harris DN, Kessler MD, et al. Sequencing of 53,831 diverse genomes from the NHLBI TOPMed Program. Nature 2021;590(7845):290–299. DOI: 10.1038/s41586-021-03205-y.

31. Das S, Forer L, Schonherr S, et al. Next-generation genotype imputation service and methods. Nat Genet 2016;48(10):1284–1287. DOI: 10.1038/ng.3656.

32. OCAC. Ovarian Cancer Association Consortium. (https://ocac.ccge.medschl.cam.ac.uk/).

33. Chenevix-Trench G, Milne RL, Antoniou AC, et al. An international initiative to identify genetic modifiers of cancer risk in BRCA1 and BRCA2 mutation carriers: the Consortium of Investigators of Modifiers of BRCA1 and BRCA2 (CIMBA). Breast Cancer Res 2007;9(2):104. DOI: 10.1186/bcr1670.

34. CIMBA. Consortium of Investigators of Modifiers of BRCA1 & BRCA2. (https://cimba.ccge.medschl.cam.ac.uk/).

35. Bycroft C, Freeman C, Petkova D, et al. The UK Biobank resource with deep phenotyping and genomic data. Nature 2018;562(7726):203–209. DOI: 10.1038/s41586-018-0579-z.

36. Biobank U. UK Biobank. (https://www.ukbiobank.ac.uk/).

37. Kurki MI, Karjalainen J, Palta P, et al. FinnGen provides genetic insights from a well-phenotyped isolated population. Nature 2023;613(7944):508–518. DOI: 10.1038/s41586-022-05473-8.

38. FinnGen. FinnGen. (https://www.finngen.fi/en).

39. Nagai A, Hirata M, Kamatani Y, et al. Overview of the BioBank Japan Project: Study design and profile. J Epidemiol 2017;27(3S):S2–S8. DOI: 10.1016/j.je.2016.12.005.

40. Japan B. BioBank Japan. (https://biobankjp.org/en/).

41. Consortium UK, Walter K, Min JL, et al. The UK10K project identifies rare variants in health and disease. Nature 2015;526(7571):82–90. DOI: 10.1038/nature14962.

42. UK10K. UK10K Project. (https://www.uk10k.org/).

43. Amos CI, Dennis J, Wang Z, et al. The OncoArray Consortium: A Network for Understanding the Genetic Architecture of Common Cancers. Cancer Epidemiol Biomarkers Prev 2017;26(1):126–135. DOI:10.1158/1055-9965.EPI-16-0106.

44. Antoniou AC, Sinilnikova OM, Simard J, et al. RAD51 135G-->C modifies breast cancer risk among BRCA2 mutation carriers: results from a combined analysis of 19 studies. Am J Hum Genet 2007;81(6):1186–200. DOI: 10.1086/522611.

45. Barnes DR, Lee A, Investigators E, kConFab I, Easton DF, Antoniou AC. Evaluation of association methods for analysing modifiers of disease risk in carriers of high-risk mutations. Genet Epidemiol 2012;36(3):274–91. DOI: 10.1002/gepi.21620.

46. Lee A, Mavaddat N, Cunningham A, et al. Enhancing the BOADICEA cancer risk prediction model to incorporate new data on RAD51C, RAD51D, BARD1 updates to tumour pathology and cancer incidence. J Med Genet 2022;59(12):1206–1218. DOI: 10.1136/jmedgenet-2022-108471.

47. Antoniou AC, Wang X, Fredericksen ZS, et al. A locus on 19p13 modifies risk of breast cancer in BRCA1 mutation carriers and is associated with hormone receptor-negative breast cancer in the general population. Nat Genet 2010;42(10):885–92. DOI: 10.1038/ng.669.

48. Willer CJ, Li Y, Abecasis GR. METAL: fast and efficient meta-analysis of genomewide association scans. Bioinformatics 2010;26(17):2190–1. DOI: 10.1093/bioinformatics/btq340.

49. Yang J, Ferreira T, Morris AP, et al. Conditional and joint multiple-SNP analysis of GWAS summary statistics identifies additional variants influencing complex traits. Nat Genet 2012;44(4):369–75, S1-3. DOI: 10.1038/ng.2213.

50. Wakefield J. A Bayesian measure of the probability of false discovery in genetic epidemiology studies. Am J Hum Genet 2007;81(2):208–27. DOI: 10.1086/519024.

51. Udler MS, Tyrer J, Easton DF. Evaluating the power to discriminate between highly correlated SNPs in genetic association studies. Genet Epidemiol 2010;34(5):463–8. DOI: 10.1002/gepi.20504.

52. Dareng EO, Tyrer JP, Barnes DR, et al. Polygenic risk modeling for prediction of epithelial ovarian cancer risk. Eur J Hum Genet 2022;30(3):349–362. DOI: 10.1038/s41431-021-00987-7.

53. Tyrer JP, Peng P, DeVries AA, Gayther SA, Jones MR, Pharoah PD. Improving on polygenic scores across complex traits using select and shrink with summary statistics. medRxiv 2022:2022.09.13.22278911. DOI: 10.1101/2022.09.13.22278911.

54. Antoniou AC, Beesley J, McGuffog L, et al. Common breast cancer susceptibility alleles and the risk of breast cancer for BRCA1 and BRCA2 mutation carriers: implications for risk prediction. Cancer Res 2010;70(23):9742–54. DOI: 10.1158/0008-5472.CAN-10-1907.

55. UK CR. Cancer Research UK: Ovarian Cancer incidence by age. (https://www.cancerresearchuk.org/health-professional/cancer-statistics/statistics-by-cancer-type/ovarian-cancer/incidence#heading-One).

56. Kuchenbaecker KB, Hopper JL, Barnes DR, et al. Risks of Breast, Ovarian, and Contralateral Breast Cancer for BRCA1 and BRCA2 Mutation Carriers. JAMA 2017;317(23):2402–2416. DOI: 10.1001/jama.2017.7112.

57. Huang L, Rosen JD, Sun Q, et al. TOP-LD: A tool to explore linkage disequilibrium with TOPMed whole-genome sequence data. Am J Hum Genet 2022;109(6):1175–1181. DOI: 10.1016/j.ajhg.2022.04.006.

58. Xiao R, Boehnke M. Quantifying and correcting for the winner’s curse in genetic association studies. Genet Epidemiol 2009;33(5):453–62. DOI: 10.1002/gepi.20398.

59. Zhang H, Ahearn TU, Lecarpentier J, et al. Genome-wide association study identifies 32 novel breast cancer susceptibility loci from overall and subtype-specific analyses. Nat Genet 2020;52(6):572–581. DOI: 10.1038/s41588-020-0609-2.

60. Stacey SN, Sulem P, Jonasdottir A, et al. A germline variant in the TP53 polyadenylation signal confers cancer susceptibility. Nat Genet 2011;43(11):1098–103. DOI: 10.1038/ng.926.

61. Stacey SN, Sulem P, Gudbjartsson DF, et al. Germline sequence variants in TGM3 and RGS22 confer risk of basal cell carcinoma. Hum Mol Genet 2014;23(11):3045–53. DOI: 10.1093/hmg/ddt671.

62. Stacey SN, Helgason H, Gudjonsson SA, et al. New basal cell carcinoma susceptibility loci. Nat Commun 2015;6:6825. DOI: 10.1038/ncomms7825.

63. Chahal HS, Wu W, Ransohoff KJ, et al. Genome-wide association study identifies 14 novel risk alleles associated with basal cell carcinoma. Nat Commun 2016;7:12510. DOI: 10.1038/ncomms12510.

64. Di Giovannantonio M, Harris BH, Zhang P, et al. Heritable genetic variants in key cancer genes link cancer risk with anthropometric traits. J Med Genet 2021;58(6):392–399. DOI: 10.1136/jmedgenet-2019-106799.

65. Melin BS, Barnholtz-Sloan JS, Wrensch MR, et al. Genome-wide association study of glioma subtypes identifies specific differences in genetic susceptibility to glioblastoma and non-glioblastoma tumors. Nat Genet 2017;49(5):789–794. DOI: 10.1038/ng.3823.

66. Egan KM, Nabors LB, Olson JJ, et al. Rare TP53 genetic variant associated with glioma risk and outcome. J Med Genet 2012;49(7):420–1. DOI: 10.1136/jmedgenet-2012-100941.

67. Enciso-Mora V, Hosking FJ, Di Stefano AL, et al. Low penetrance susceptibility to glioma is caused by the TP53 variant rs78378222. Br J Cancer 2013;108(10):2178–85. DOI: 10.1038/bjc.2013.155.

68. Wang Z, Rajaraman P, Melin BS, et al. Further Confirmation of Germline Glioma Risk Variant rs78378222 in TP53 and Its Implication in Tumor Tissues via Integrative Analysis of TCGA Data. Hum Mutat 2015;36(7):684–8. DOI: 10.1002/humu.22799.

69. Conti DV, Darst BF, Moss LC, et al. Trans-ancestry genome-wide association meta-analysis of prostate cancer identifies new susceptibility loci and informs genetic risk prediction. Nat Genet 2021;53(1):65–75. DOI: 10.1038/s41588-020-00748-0.

70. Li Y, Gordon MW, Xu-Monette ZY, et al. Single nucleotide variation in the TP53 3’ untranslated region in diffuse large B-cell lymphoma treated with rituximab-CHOP: a report from the International DLBCL Rituximab-CHOP Consortium Program. Blood 2013;121(22):4529–40. DOI: 10.1182/blood-2012-12-471722.

71. Bode AM, Dong Z. Post-translational modification of p53 in tumorigenesis. Nat Rev Cancer 2004;4(10):793–805. DOI: 10.1038/nrc1455.

72. Olivier M, Hussain SP, Caron de Fromentel C, Hainaut P, Harris CC. TP53 mutation spectra and load: a tool for generating hypotheses on the etiology of cancer. IARC Sci Publ 2004(157):247–70. (https://www.ncbi.nlm.nih.gov/pubmed/15055300).

73. Levine AJ. p53, the cellular gatekeeper for growth and division. Cell 1997;88(3):323–31. DOI: 10.1016/s0092-8674(00)81871-1.

74. Vogelstein B, Lane D, Levine AJ. Surfing the p53 network. Nature 2000;408(6810):307–10. DOI: 10.1038/35042675.

75. Zhang P, Kitchen-Smith I, Xiong L, et al. Germline and Somatic Genetic Variants in the p53 Pathway Interact to Affect Cancer Risk, Progression, and Drug Response. Cancer Res 2021;81(7):1667–1680. DOI: 10.1158/0008-5472.CAN-20-0177.

76. Schildkraut JM, Goode EL, Clyde MA, et al. Single nucleotide polymorphisms in the TP53 region and susceptibility to invasive epithelial ovarian cancer. Cancer Res 2009;69(6):2349–57. DOI: 10.1158/0008-5472.CAN-08-2902.

77. GTEx. Genotype-Tissue Expression project. (https://gtexportal.org/home/).

78. Urbanek M, Legro RS, Driscoll DA, et al. Thirty-seven candidate genes for polycystic ovary syndrome: strongest evidence for linkage is with follistatin. Proc Natl Acad Sci U S A 1999;96(15):8573–8. DOI: 10.1073/pnas.96.15.8573.

79. Frandsen CLB, Svendsen PF, Nohr B, et al. Risk of epithelial ovarian tumors among women with polycystic ovary syndrome: A nationwide population-based cohort study. Int J Cancer 2023;153(5):958–968. DOI: 10.1002/ijc.34574.

80. Harris HR, Cushing-Haugen KL, Webb PM, et al. Association between genetically predicted polycystic ovary syndrome and ovarian cancer: a Mendelian randomization study. Int J Epidemiol 2019;48(3):822–830. DOI: 10.1093/ije/dyz113.

81. Manichaikul A, Peres LC, Wang XQ, et al. Identification of novel epithelial ovarian cancer loci in women of African ancestry. Int J Cancer 2020;146(11):2987–2998. DOI: 10.1002/ijc.32653.

82. Lin CH, Vu JP, Yang CY, et al. Glutamate-cysteine ligase catalytic subunit as a therapeutic target in acute myeloid leukemia and solid tumors. Am J Cancer Res 2021;11(6):2911–2927. (https://www.ncbi.nlm.nih.gov/pubmed/34249435).

83. Ogiwara H, Takahashi K, Sasaki M, et al. Targeting the Vulnerability of Glutathione Metabolism in ARID1A-Deficient Cancers. Cancer Cell 2019;35(2):177–190 e8. DOI: 10.1016/j.ccell.2018.12.009.

84. Wu RC, Wang TL, Shih Ie M. The emerging roles of ARID1A in tumor suppression. Cancer Biol Ther 2014;15(6):655–64. DOI: 10.4161/cbt.28411.

85. Li W, Lv D, Yao J, et al. A pan-cancer analysis reveals the diagnostic and prognostic role of CDCA2 in low-grade glioma. PLOS ONE 2023;18(9):e0291024. DOI: 10.1371/journal.pone.0291024.

86. Veeriah S, Brennan C, Meng S, et al. The tyrosine phosphatase PTPRD is a tumor suppressor that is frequently inactivated and mutated in glioblastoma and other human cancers. Proc Natl Acad Sci U S A 2009;106(23):9435–40. DOI: 10.1073/pnas.0900571106.

87. Sherry ST, Ward M, Sirotkin K. dbSNP-database for single nucleotide polymorphisms and other classes of minor genetic variation. Genome Res 1999;9(8):677–9. (https://www.ncbi.nlm.nih.gov/pubmed/10447503).

88. Rafnar T, Gunnarsson B, Stefansson OA, et al. Variants associating with uterine leiomyoma highlight genetic background shared by various cancers and hormone-related traits. Nat Commun 2018;9(1):3636. DOI: 10.1038/s41467-018-05428-6.

89. Harris HR, Peres LC, Johnson CE, et al. Racial Differences in the Association of Endometriosis and Uterine Leiomyomas With the Risk of Ovarian Cancer. Obstet Gynecol 2023;141(6):1124–1138. DOI: 10.1097/AOG.0000000000005191.

90. Broggini M, Buraggi G, Brenna A, et al. Cell cycle-related phosphatases CDC25A and B expression correlates with survival in ovarian cancer patients. Anticancer Res 2000;20(6C):4835–40. (https://www.ncbi.nlm.nih.gov/pubmed/11205229).

91. Schraml P, Bucher C, Bissig H, et al. Cyclin E overexpression and amplification in human tumours. J Pathol 2003;200(3):375–82. DOI: 10.1002/path.1356.

92. 92. Cancer Genome Atlas Research N. Integrated genomic analyses of ovarian carcinoma. Nature 2011;474(7353):609–15. DOI: 10.1038/nature10166.

93. Goundiam O, Gestraud P, Popova T, et al. Histo-genomic stratification reveals the frequent amplification/overexpression of CCNE1 and BRD4 genes in non-BRCAness high grade ovarian carcinoma. Int J Cancer 2015;137(8):1890–900. DOI: 10.1002/ijc.29568.

94. Kang EY, Weir A, Meagher NS, et al. CCNE1 and survival of patients with tubo-ovarian high-grade serous carcinoma: An Ovarian Tumor Tissue Analysis consortium study. Cancer 2023;129(5):697–713. DOI: 10.1002/cncr.34582.

95. 95. Cancer Genome Atlas N. Comprehensive molecular portraits of human breast tumours. Nature 2012;490(7418):61–70. DOI: 10.1038/nature11412.

96. Curtis C, Shah SP, Chin SF, et al. The genomic and transcriptomic architecture of 2,000 breast tumours reveals novel subgroups. Nature 2012;486(7403):346–52. DOI: 10.1038/nature10983.

97. Jiang YZ, Ma D, Suo C, et al. Genomic and Transcriptomic Landscape of Triple-Negative Breast Cancers: Subtypes and Treatment Strategies. Cancer Cell 2019;35(3):428–440 e5. DOI: 10.1016/j.ccell.2019.02.001.

98. Jones RM, Mortusewicz O, Afzal I, et al. Increased replication initiation and conflicts with transcription underlie Cyclin E-induced replication stress. Oncogene 2013;32(32):3744–53. DOI: 10.1038/onc.2012.387.

99. Gallo D, Young JTF, Fourtounis J, et al. CCNE1 amplification is synthetic lethal with PKMYT1 kinase inhibition. Nature 2022;604(7907):749–756. DOI: 10.1038/s41586-022-04638-9.

100. Lee A, Yang X, Tyrer J, et al. Comprehensive epithelial tubo-ovarian cancer risk prediction model incorporating genetic and epidemiological risk factors. J Med Genet 2022;59(7):632–643. DOI: 10.1136/jmedgenet-2021-107904.

101. Brnich SE, Abou Tayoun AN, Couch FJ, et al. Recommendations for application of the functional evidence PS3/BS3 criterion using the ACMG/AMP sequence variant interpretation framework. Genome Med 2019;12(1):3. DOI: 10.1186/s13073-019-0690-2.

102. Staley JR, Blackshaw J, Kamat MA, et al. PhenoScanner: a database of human genotype-phenotype associations. Bioinformatics 2016;32(20):3207–3209. DOI: 10.1093/bioinformatics/btw373.

103. Kamat MA, Blackshaw JA, Young R, et al. PhenoScanner V2: an expanded tool for searching human genotype-phenotype associations. Bioinformatics 2019;35(22):4851–4853. DOI: 10.1093/bioinformatics/btz469.

104. PhenoScanner. PhenoScanner V2: A database of human genotype-phenotype associations. (http://www.phenoscanner.medschl.cam.ac.uk/).

105. PheWeb. PheWeb. (https://pheweb.org/UKB-TOPMed/).

106. Vosa U, Claringbould A, Westra HJ, et al. Large-scale cis- and trans-eQTL analyses identify thousands of genetic loci and polygenic scores that regulate blood gene expression. Nat Genet 2021;53(9):1300–1310. DOI: 10.1038/s41588-021-00913-z.

107. eQTLGen. eQTLGen. (https://www.eqtlgen.org/).

