## Supplementary Material for "Large-scale genome-wide association study of 398,238 women unveils seven novel loci associated with high-grade serous epithelial ovarian cancer risk"

### SUPPLEMENTARY METHODS

*Genotyping*

Genotyping of OCAC and CIMBA samples were performed on one of two custom single nucleotide polymorphism (SNP) genotyping arrays, the iCOGS[1,2] array or OncoArray[3-5]. The iCOGS array included approximately 210,000 SNPs that were selected for previous evidence of association with breast, ovarian and prostate cancer. The OncoArray is a custom genotyping chip consisting of approximately 533,000 SNPs, approximately half of which is a GWAS backbone that tags common SNPs. A standard quality control (QC) process was applied, including assessment of SNP call rate, allele frequency, genotyping intensity clustering, Hardy-Weinberg equilibrium, and SNP concordance from duplicated samples[4].

*Principal components analysis*

Principal components (PCs) for OncoArray data were calculated using 33,661 uncorrelated ($r^2$<0.10) common (MAF>0.05) SNPs. Calculations were performed using a custom program, available at http://ccge.medschl.cam.ac.uk/software/pccalc/. Details of PC calculations for the other genotype data has been previously described elsewhere[1,6].

*Ancestry analysis*

Ancestry for OCAC data was calculated using the FastPop software[7]. Women with >80% European ancestry were retained for statistical analyses. For CIMBA data, 33,661 common uncorrelated SNPs (the same set used to calculate the PCs, described above) were used to calculate kinship coefficients between all CIMBA participants and 267 HapMap samples (CHD, JPT, YRI and CEU ancestries). These kinship coefficients were converted to distances and then underwent multidimensional scaling. Using the top two PCs, the proportion of European ancestry for each participant. Women with >27% non-European ancestry were excluded to ensure women with Ashkenazi Jewish ancestry were retained for statistical analyses.

*Imputation to the TOPMed reference panel*

Genotyped samples were imputed using the Michigan Imputation Server[8,9] to the TOPMed imputation panel[10] with 97,256 samples (Version R2 on GRC38). Phasing was performed with Eagle2[11] and imputation with Minimac[12,13]. Prior to imputation, variants were excluded from the genotype files using the following criteria: common variants with a call rate below 95%; rare variants (MAF<1%) with a call rate below 98%; variants not in Hardy–Weinberg equilibrium ($P<1x10^{-7}$ in controls, or $P<1x10^{-12}$ in cases); variants with poor cluster plots. We then used a script (HRC-1000G-check-bim-v4.3.0.pl available at https://www.well.ox.ac.uk/~wrayner/tools/) to remove variants not on the TOPMed reference panel or align them to the correct strand. This tool excluded variants where the genotyped frequency differed from the panel by more than 0.20.

Samples were randomised into batches of less than 25,000 to meet the maximum sample requirement of the imputation server[8-10] and the same list of variants was included for each batch of genotypes.

*Sensitivity analyses for genome-wide statistically significant associations*

Variants exhibiting genome-wide statistically significant associations were re-evaluated to minimise spurious associations. The associations in OCAC and UKBB were reanalysed by pooling all individual participant level data. These models accounted for specific genotyping project or study, and incorporated 29 principal components derived from all genotyping projects (set to 0 if not present for a specific study/project). The UKBB data were adjusted for age, which was set to 0 for the OCAC data. Any variant with imputation accuracy $r^2<0.30$ within a panel were considered missing for that particular panel.

*Polygenic models*

We tested a total of 1,102 PGMs, each of which was a combination of the S4 model hyperparameters. There were 562 models taking variants with P-value/$r^2<0.02$ (resulting in ~64k variants), and 540 models taking variants with P-value/$r^2<0.15$ (resulting in ~394k variants). Models with P-value/$r^2<0.15$ did not substantially improve PGS performance to compensate for the far larger number of selected variants.

*Candidate clinical PGS development*

We first selected the single nucleotide variants that were genotyped on the OncoArray, since they are reliably genotyped, and many had been chosen for their relevance to ovarian cancer. We then selected a subset of genotyped variants, based on “relative importance”, where the relative importance of each variant is approximately proportional to $p$*(1-$p$)*$\beta^2$, where β is the log-RR and $p$ is the minor allele frequency for that variant. We ranked each variant based on relative importance and selected the top *N* desired variants.

### SUPPLEMENTARY RESULTS

*S4 PGM hyperparameters*

The best performing PGS contained 64,518 variants, that had S4 hyperparameters: $\alpha = 0.1$, $\beta = 0.8$ and $\varphi = 8\times10^{-8}$. $\alpha$ controls the degree of shrinkage towards zero for the model coefficients. A smaller $\alpha$ means more shrinkage, which implies more sparsity and less variance. $\beta$ controls the amount of shrinkage for extreme values of the model coefficients. A larger $\beta$ means more shrinkage for extreme values. $\varphi$ controls the overall scale of the shrinkage in the model. A larger $\varphi$ means a larger scale, which implies more variability and less shrinkage for all coefficients.

**ETHICS STATEMENT**

All study participants provided written informed consent and participated in research studies at the host institute under ethically approved protocols.

All study participants provided written informed consent and participated in research or clinical studies at the host institute under ethically approved protocols. The studies and their approving institutes are: Australian site of the Breast Cancer Family Registry (BCFR-AU) - The University of Melbourne Health Sciences Human Ethics Sub-Committee; Northern California site of the Breast Cancer Family Registry (BCFR-NC) - Northern California Cancer Center Institutional Review Board; New York site of the Breast Cancer Family Registry (BCFR-NY) - Columbia University Medical Center Institutional Review Board; Ontario site of the Breast Cancer Family Registry (BCFR-ON) - Mount Sinai Hospital Research Ethics Board; Philadelphia site of the Breast Cancer Family Registry (BCFR-PA) - Institutional Review Board Fox Chase Cancer Center; Utah site of the Breast Cancer Family Registry (BCFR-UT) - Institutional Review Board University of Utah; Baltic Familial Breast and Ovarian Cancer Consortium (BFBOCC) - Centrālā medicīnas ētikas Komiteja; Lietuvos Bioetikos Komitetas; BRCA-gene mutations and breast cancer in South African women (BMBSA) - University of Pretoria and Pretoria Academic Hospitals Ethics Committee; Beckman Research Institute of the City of Hope (BRICOH) - City of Hope Medical Center Institutional Review Board; Copenhagen Breast Cancer Study (CBCS) - De Videnskabsetiske Komiteer I Region Hovedsladen; Spanish National Cancer Centre (CNIO) - Instituto de Salud Carlos III Comité de Bioética y Bienestar Animal; City of Hope Cancer Center (COH) - City of Hope Institutional Review Board; CONsorzio Studi ITaliani sui Tumori Ereditari Alla Mammella (CONSIT TEAM) - Comitato Etico Indipendente della Fondazione IRCCS "Istituto Nazionale dei

Tumori"; National Centre for Scientific Research Demokritos (DEMOKRITOS) - Bioethics committee of NCSR ‘‘Demokritos’’, 240/EHΔ/11.3; National Centre for Scientific Research Demokritos (DEMOKRITOS) - Papageorgiou Hospital Ethics Committee; Dana Farber Cancer Institute (DFCI) - Dana Farber Cancer Institute Institutional Review Board; Deutsches Krebsforschungszentrum (DKFZ) - Ethik-Kommission des Klinikums der Universität; Deutsches Krebsforschungszentrum (DKFZ) - Hospital Universitario de San Ignacio Comité de Investigaciones y Etica; Deutsches Krebsforschungszentrum (DKFZ) - Shaukat Khanum Memorial Cancer Hospital and Research Centre Institutional Review Board; Epidemiological study of BRCA1 and BRCA2 mutation carriers (EMBRACE) - Anglia & Oxford MREC; Fox Chase Cancer Center (FCCC) - Institutional Review Board Fox Chase Cancer Center; Fundación Pública Galega de Medicina Xenómica - Comite Autonomico de Etica da Investigacion de Galicia; German Consortium of Hereditary Breast and Ovarian Cancer (GC-HBOC) - Ethik-Kommission der Medizinischen Fakultät der Universät zu Köln; Genetic Modifiers of cancer risk in BRCA1/2 mutation carriers (GEMO) - Comité consultatif sur le traitement de l'information en matière de recherche dans le domaine de la santé; Georgetown University (GEORGETOWN) - MedStar Research Institute - Georgetown University Oncology Institutional Review Board; Ghent University Hospital (G-FAST) - Universitair Ziekenhuis Gent - Ethics Committee; Hospital Clinico San Carlos (HCSC) - Comité Ético de Investigación Clínia Hospital Clínico San Carlos; Helsinki Breast Cancer Study (HEBCS) - Helsingin ja uudenmaan sairaanhoitopiiri (Helsinki University Central Hospital ethics committee); HEreditary Breast and Ovarian study Netherlands (HEBON) - Protocol Toetsingscommissie van het Nederlands Kanker Instituut/Antoni van Leeuwenhoek Ziekenhuis; Molecular Genetic Studies of Breast- and Ovarian Cancer in Hungary

(HUNBOCS) - Institutional Review Board of the Hungarian National Institute of Oncology; University Hospital Vall d'Hebron (HVH) - The Hospital Universitario Vall d'Hebron Clinical Research Ethics Committee; Institut Català d'Oncologia (ICO) - Catalan Institute of Oncology Institutional Review Board; International Hereditary Cancer Centre (IHCC) - Komisji Bioetycznej Pomorskiej Akademii Medycznej (Pomeranian Medical University Bioethics Committee); Iceland Landspitali - University Hospital (ILUH) - Vísindasiđanefnd National Boethics Committee; Interdisciplinary Health Research International Team Breast Cancer Susceptibility (INHERIT) - Comité d'éthique de la recherche du Centre Hospitalier Universitaire de Québec; Istituto Oncologico Veneto Hereditary Breast and Ovarian Cancer Study (IOVHBOCS) - Centro Oncologico Regionale Azienda Ospedale Di Padova Comitato Etico; Portuguese Oncology Institute-Porto Breast Cancer Study - COMISSÃO DE ÉTICA PARA A SAÚDE (CES) ; Kathleen Cuningham Foundation Consortium for Research into Familial Breast Cancer (KCONFAB) - Queensland Institute of Medical Research - Human Research Ethics Committee; Kathleen Cuningham Foundation Consortium for Research into Familial Breast Cancer (KCONFAB) - Peter MacCallum Cancer Centre Ethics Committee; University of Kansas Medical Center(KUMC) - The University of Kansas Medical Center Human Research Protection Program; Mayo Clinic (MAYO) - Mayo Clinic Institutional Review Boards; McGill University (MCGILL) - McGill Faculty of Medicine Institutional Review Board; Modifier Study of Quantitative Effects on Disease (MOD-SQUAD) - Mayo Clinic Institutional Review Boards; Memorial Sloane Kettering Cancer Center (MSKCC) - Human Biospecimen Utilization Committee; Memorial Sloane Kettering Cancer Center (MSKCC) - Memorial Sloan-Kettering Cancer Center IRB; General Hospital Vienna (MUV) - Ethikkommission der Medizinischen Universität Wien; Women’s

College Research Institute Hereditary Breast and Ovarian Cancer Study - University of Toronto Health Sciences Review Ethics Board; National Cancer Institute (NCI) - NIH Ethics Office; National Israeli Cancer Control Center (NICCC) - Carmel Medical Center Institutional Review Board (Helsinki Committee); N.N. Petrov Institute of Oncology (NNPIO) - N.N. Petrov Institional Ethical Committee; NorthShore University HealthSystem (NORTHSHORE) - Institutional Review Board of NorthShore University HealthSystem; NRG Oncology (NRG_ONCOLOGY) - Cancer Prevention and Control Protocol Review Committee; Ontario Cancer Genetics Network (OCGN) - University Health Network Research Ethics Board; The Ohio State University Comprehensive Cancer Center (MACBRCA) - The Ohio State University Cancer Institutional Review Board; Odense University Hospital (OUH) - Den Videnskabsetiske Komité for Region Syddanmark; Pisa Breast Cancer Study (PBCS) - Azienda Ospedaliera Pisana Comitato Etico per lo studio del farmaco sull'uomo; Sheba Medical Centre - Chaim Sheba Medical Center IRB; Swedish Breast Cancer Study (SWE-BRCA) - Regionala Etikprövningsnämnden Stockholm; University of Chicago (UCHICAGO) - The University of Chicago Biological Sciences Division. Institutional Review Board (BSD IRB); University of California Los Angeles (UCLA) - UCLA Institutional Review Board (UCLA IRB); University of California San Francisco (UCSF) - Human Research Protection Program Institutional Review Board (IRB); UK and Gilda Radner Familial Ovarian Cancer Registries (UKGRFOCR) - Roswell Park Cancer Institute IRB; UK and Gilda Radner Familial Ovarian Cancer Registries (UKGRFOCR) - Cambridge Local Research Ethics Committee; University of Pennsylvania (UPENN) - University of Pennsylvania Institutional Review Board; Cancer Family Registry University of Pittsburg (UPITT) - University of Pittsburgh Institutional Review Board; University of Texas MD Anderson Cancer Center

(UTMDACC) - University of Texas MD Anderson Cancer Center Office of Protocol Research Institutional Review Board; Victorian Familial Cancer Trials Group (VFCTG) - Peter MacCallum Cancer Centre Ethics Committee; Women's Cancer Program at Cedars-Sinai Medical Center (WCP) - (Cedars-Sinai Medical Center) CSMC Institutional Review Board.

### REFERENCES


1. Couch FJ, Wang X, McGuffog L, et al. Genome-wide association study in BRCA1 mutation carriers identifies novel loci associated with breast and ovarian cancer risk. PLoS Genet 2013;9(3):e1003212. DOI: 10.1371/journal.pgen.1003212.
2. Gaudet MM, Kuchenbaecker KB, Vijai J, et al. Identification of a BRCA2-specific modifier locus at 6p24 related to breast cancer risk. PLoS Genet 2013;9(3):e1003173. DOI: 10.1371/journal.pgen.1003173.
3. Amos CI, Dennis J, Wang Z, et al. The OncoArray Consortium: A Network for Understanding the Genetic Architecture of Common Cancers. Cancer Epidemiol Biomarkers Prev 2017;26(1):126-135. DOI: 10.1158/1055-9965.EPI-16-0106.
4. Milne RL, Kuchenbaecker KB, Michailidou K, et al. Identification of ten variants associated with risk of estrogen-receptor-negative breast cancer. Nat Genet 2017;49(12):1767-1778. DOI: 10.1038/ng.3785.
5. Phelan CM, Kuchenbaecker KB, Tyrer JP, et al. Identification of 12 new susceptibility loci for different histotypes of epithelial ovarian cancer. Nat Genet 2017;49(5):680-691. DOI: 10.1038/ng.3826.
6. Pharoah PD, Tsai YY, Ramus SJ, et al. GWAS meta-analysis and replication identifies three new susceptibility loci for ovarian cancer. Nat Genet 2013;45(4):362-70, 370e1-2. DOI: 10.1038/ng.2564.
7. Li Y, Byun J, Cai G, et al. FastPop: a rapid principal component derived method to infer intercontinental ancestry using genetic data. BMC Bioinformatics 2016;17:122. DOI: 10.1186/s12859-016-0965-1.

8. Das S, Forer L, Schonherr S, et al. Next-generation genotype imputation service and methods. Nat Genet 2016;48(10):1284-1287. DOI: 10.1038/ng.3656.

9. TOPMed. TOPMed Imputation Server. (https://imputation.biodatacatalyst.nhlbi.nih.gov/).

10. Taliun D, Harris DN, Kessler MD, et al. Sequencing of 53,831 diverse genomes from the NHLBI TOPMed Program. Nature 2021;590(7845):290-299. DOI: 10.1038/s41586-021-03205-y.

11. Loh PR, Danecek P, Palamara PF, et al. Reference-based phasing using the Haplotype Reference Consortium panel. Nat Genet 2016;48(11):1443-1448. DOI: 10.1038/ng.3679.

12. Howie B, Fuchsberger C, Stephens M, Marchini J, Abecasis GR. Fast and accurate genotype imputation in genome-wide association studies through pre-phasing. Nat Genet 2012;44(8):955-9. DOI: 10.1038/ng.2354.

13. Fuchsberger C, Abecasis GR, Hinds DA. minimac2: faster genotype imputation. Bioinformatics 2015;31(5):782-4. DOI: 10.1093/bioinformatics/btu704.
